# IN-HOSPITAL CONTINUATION WITH ANGIOTENSIN RECEPTOR BLOCKERS IS ASSOCIATED WITH A LOWER MORTALITY RATE THAN CONTINUATION WITH ANGIOTENSIN CONVERTING ENZYME INHIBITORS IN COVID-19 PATIENTS: A RETROSPECTIVE COHORT STUDY

**DOI:** 10.1101/2021.02.01.21250853

**Authors:** Francisco J. de Abajo, Antonio Rodríguez-Miguel, Sara Rodríguez-Martín, Victoria Lerma, Alberto García-Lledó, on behalf of MED-ACE2-COVID19 Study Group

**Author notes:** **Author for correspondence:** Prof. Francisco J. de Abajo, Clinical Pharmacology Unit, University Hospital Principe de Asturias, Department of Biomedical Sciences, University of Alcalá (IRYCIS), (ext 2607) - 91 885 25 93. **MED-ACE2-COVID19 Study Group**: *Hospital Universitario Príncipe de Asturias*: F J de Abajo, A Rodríguez-Miguel, S Rodríguez-Martín, V Lerma, A García-Lledó, D Barreira-Hernández; D Rodríguez-Puyol; *Hospital Universitario de Getafe*: O Laosa, L Pedraza, L Rodríguez-Mañas; *Hospital Universitario Ramón y Cajal*: M Aguilar, I de Pablo, MA Gálvez; *Hospital Central de la Defensa Gómez Ulla*: A García-Luque, M Puerro; RM Aparicio, V García-Rosado, C Gutiérrez-Ortega; *Hospital Clínico San Carlos*: L Laredo, E González-Rojano, C Pérez, A Ascaso, C Elvira; *Hospital Universitario de La Princesa*: G Mejía-Abril, P Zubiaur, E Santos-Molina, E Pintos-Sánchez, M Navares-Gómez; F Abad-Santos; *Hospital Universitario Puerta de Hierro-Majadahonda*: G A Centeno, A Sancho-Lopez, C Payares-Herrera, E Diago-Sempere.

## Abstract

**Background:** Several studies have reported a reduced risk of death associated with the inpatient use of angiotensin receptor blockers (ARBs) and angiotensin converting enzyme inhibitors (ACEIs) in COVID-19 patients, but have been criticized for incurring in several types of bias. Also, most studies have pooled ACEIs and ARBs as if they were a unique group, overlooking their pharmacological differences. We aimed to assess whether the in-hospital continuation of ARBs and ACEIs, in regular users of these drugs, was associated with a reduced risk of death as compared to their discontinuation and also to compare head-to-head ARBs with ACEIs.

**Methods:** Adult patients with a PCR-confirmed diagnosis of COVID-19 requiring admission during March, 2020 were consecutively selected from 7 hospitals in Madrid, Spain. Among them, we identified outpatient users of ACEIs/ARBs and divided them in two cohorts depending on treatment discontinuation/continuation at admission. Then, they were followed-up until discharge or in-hospital death. An intention-to-treat survival analysis was carried out and hazard ratios (HRs) and their 95%CI were computed through a Cox regression model adjusted for propensity scores of discontinuation and controlled by potential mediators.

**Results:** Out of 625 ACEI/ARB users, 340(54.4%) discontinued treatment. The in-hospital mortality rates were 27.6% and 27.7% in discontinuation and continuation cohorts, respectively (HR=1.01; 95%CI:0.70-1.46). No difference in mortality was observed between ARB and ACEI discontinuation (28.6% vs. 27.1%, respectively), while a significantly lower mortality rate was found among patients who continued with ARBs (20.8%,N=125) as compared to those who continued with ACEIs (33.1%,N=136; p=0.03). The head-to-head comparison (ARB vs. ACEI continuation) yielded an adjusted HR of 0.52 (95%CI:0.29-0.93), being especially notorious among males (HR=0.34; 95%CI:0.12-0.93), subjects older than 74 years (HR=0.46; 95%CI:0.25-0.85), and patients with obesity (HR=0.22; 95%CI:0.05-0.94), diabetes (HR=0.36; 95%CI:0.13-0.97) and heart failure (HR=0.12; 95%CI:0.03-0.97).

**Conclusions:** Among regular users of ARBs admitted for COVID-19, the in-hospital continuation with them was associated with an improved survival, while this was not observed with ACEIs. Regular users of ARBs should continue with this treatment if admitted for COVID-19, unless medically contraindicated. In admitted ACEI users, a switching to ARBs should be considered, especially among high-risk patients.

**GRAPHICAL ABSTRACT:** 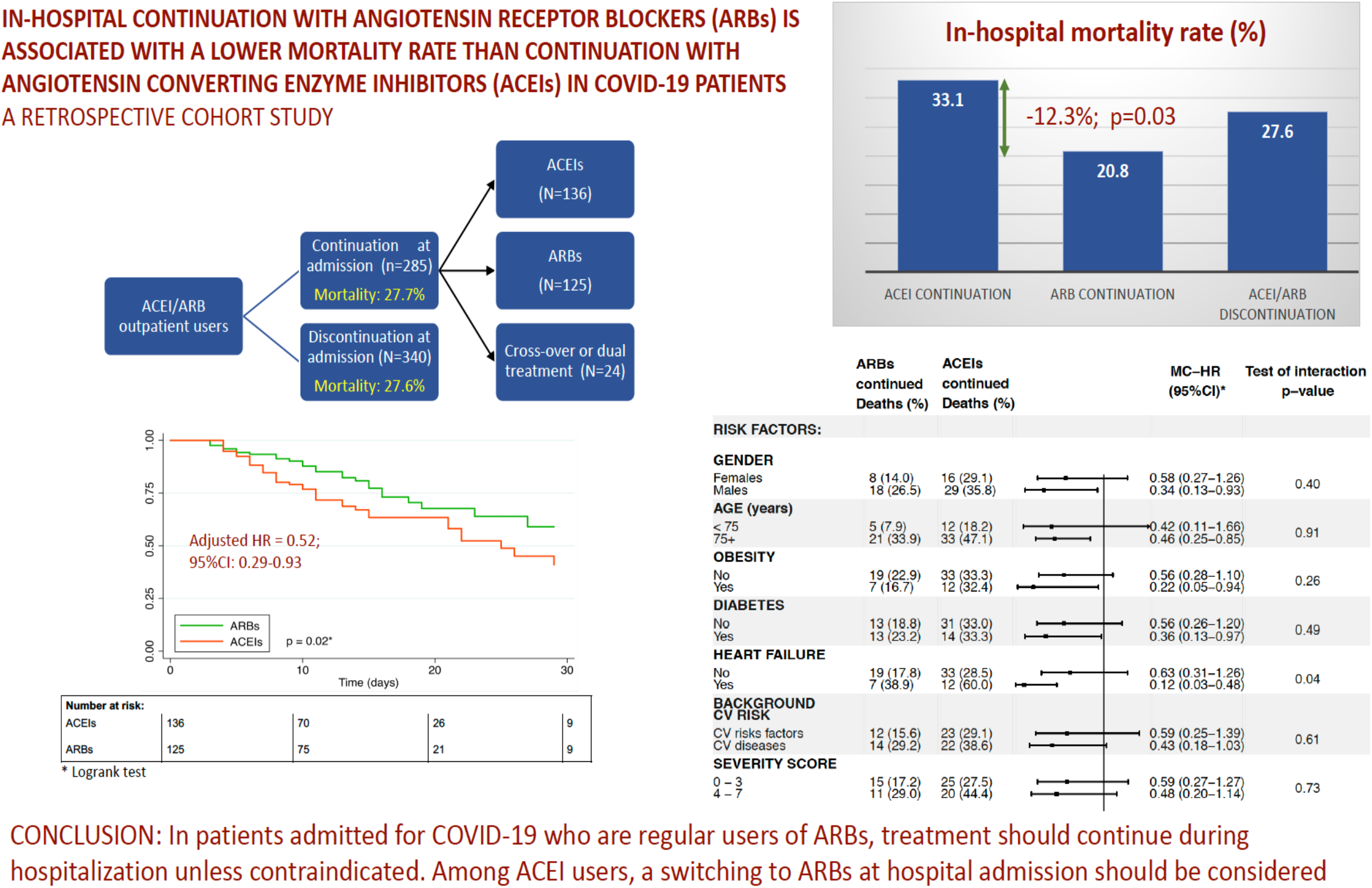

## INTRODUCTION

In mid-March, at the start of the first wave of COVID-19 pandemic in Europe, the hypothesis that the renin-angiotensin system inhibitors (RASIs), including angiotensin receptor blockers (ARBs) and angiotensin converting enzyme inhibitors (ACEIs), increased the risk and/or severity of the disease [1-3] was widely spread. Consequently, many hospitals and clinicians adopted the “precautionary measure” to discontinue these drugs from patients who regularly used them. Promptly, in the first weeks of May, three large epidemiological studies were published supporting the lack of association between the outpatient use of RASIs and risk of COVID-19 [4-6]. Later on, a plethora of studies and meta-analyses were published [7,8] reaching the same conclusion, which provide reassurance on the safety of these drugs. Yet, the extent of RASI discontinuation at hospital admission during the first wave of the pandemic and, importantly, its impact on health outcomes have been scarcely studied [9-12].

The downregulation of angiotensin-converting enzyme type 2 (ACE2), as resulted from the SARS-CoV-2 endocytosis, has been postulated to play a key role in the progression of COVID-19 to severe forms [13]. In physiological conditions, the ACE1-angiotensin II-AT1R axis (the classical RAS) is counter-regulated by the ACE2-Angiotensin (1-7)-MasR axis. Thus, when the latter weakens, angiotensin II is unopposed and its vasoconstrictor, pro-inflammatory and pro-thrombotic actions may contribute to the pathophysiology of severe COVID-19 [13-15]. In this context, it is conceivable that treatment with RASIs in COVID-19 inpatients could compensate the ACE1/ACE2 imbalance provoked by the SARS-CoV-2 and produce a net beneficial effect. According to this, several observational studies have reported a protective effect of inpatient use of RASIs on mortality as compared to non-use (or non-RASI use) in COVID-19 patients [9-12]. However, such studies have been criticized for incurring in several types of bias [16,17]. Recently, two randomized clinical trials have been published [18,19] reporting no difference in mortality between discontinuation and continuation arms. However, these trials and most observational studies have pooled ACEIs and ARBs and analyzed in a unique group, overlooking that they have different pharmacological actions [20] that could lead to distinct clinical effects [20], particularly in COVID-19 patients [15]. In this sense, no study has carried out a head-to-head (ARBs vs. ACEIs) comparison in COVID-19 patients admitted to hospital.

The present research was aimed: 1) to quantify the magnitude of RASI discontinuation at admission in seven hospitals from the Autonomous Community of Madrid, Spain; 2) to compare in real-life conditions the in-hospital mortality in patients in whom ACEIs or ARBs were discontinued with those in whom these drugs were continued; and 3) to perform a head-to-head comparison between in-hospital use of ACEIs and ARBs regarding mortality in admitted patients for COVID-19.

## PATIENTS AND METHODS

### Study design, subject selection and follow-up

We collected information from patients aged 18 years or older admitted to hospital from March 1, 2020 to March 31, 2020, with a diagnosis of COVID-19 confirmed by RT-PCR. Seven hospitals of the Autonomous Community of Madrid (Spain) took part. According to drug exposure in the month prior to admission, patients were classified in three study groups: 1) users of RASIs; 2) users of non-RASI antihypertensive drugs; and 3) non-users of antihypertensive drugs. For the present study, only RASI users were considered. Among them, we excluded those in whom the continuation or discontinuation of RASI treatment could not be properly assessed at admission, including patients transferred to another hospital from the emergency department (ED) and patients who presented the outcome (death or admission to the intensive care Unit; ICU) or were discharged within the first 3 days of hospital admission. Hence, eligible patients had to survive and be outcome-free in a hospital ward (excluding ICU) at least during the first 3 days since admission to the ED. Then, they were subdivided in two closed cohorts: 1) *Continuation cohort:* patients in whom RASI prescriptions were recorded in at least 2 of the first 3 days since ED admission (including switching from one RASI to another); and 2) *Discontinuation cohort*: patients in whom no prescription of RASI was recorded in the first 3 days since ED admission. When there was a sole prescription of RASIs in the first 3 days, the intention-to-discontinue was considered uncertain and these patients were not included in the main analysis; however, we carried out two sensitivity analyses in which these patients were re-classified (see “Sensitivity analyses”). Both cohorts were then followed-up until discharge or in-hospital death (any cause), recording any ICU admission. The date of admission to the ED was considered the index date for the follow-up, so the above definitions assume an immortal time of 3 days in both continuation and discontinuation cohorts (avoiding this way a bias).

### Sources of information and data collection

The information on co-morbidities and drug exposure before admission was extracted from electronic primary healthcare records, as described in detail elsewhere in a previous study [6]. The information on disease severity at admission and its clinical evolution (death, discharge, ICU admission and in-hospital treatment received) was retrieved from hospital medical records. All data extracted were anonymized and included in *ad hoc* case report forms in each participating hospital, then sent out to the coordinating center, where a data quality control was undertaken.

### Baseline co-morbidities and outpatient treatments

The presence of the following baseline co-morbidities were recorded at index date: antecedents of hypertension, dyslipidemia (recorded as such or when there was at least one prescription of a lipid-lowering drug), diabetes (recorded as such or when there was at least one prescription of a glucose-lowering drug), ischemic heart disease, atrial fibrillation, heart failure, thromboembolic disease, cerebrovascular accident (including stroke and transient ischemic accident), asthma, chronic obstructive pulmonary disease (COPD), chronic renal failure and cancer (past and active). We also collected information on obesity (defined as a body mass index -BMI-≥ 30 kg/m^2^), smoking (current smoker, past smoker, non-smoker or not recorded) and the outpatient use of calcium channel blockers (CCBs), beta-blocking agents, alpha-adrenoceptor antagonists with cardiovascular (CV) indications, high-ceiling diuretics, low-ceiling diuretics, antagonists of mineralocorticoid receptor (AMRs), lipid-lowering drugs, glucose-lowering drugs, antiplatelet drugs, oral anticoagulants, nonsteroidal anti-inflammatory drugs (NSAIDS), systemic corticosteroids and non-opioid analgesics (paracetamol and metamizole).

### Disease severity

To characterize the severity of COVID-19 at admission we collected information on the presence of pneumonia, hypoxemia (defined as oxygen saturation ≤90% at rest breathing ambient air, or a PaO2/FiO2 ratio ≤300 mm Hg), lymphopenia, and abnormal values of five inflammatory biomarkers (according to the reference values of each hospital laboratory), when available: C-protein reactive (CPR), procalcitonin, troponin, D-dimer, and N-terminal type B natriuretic propeptide (NT-pro-BNP)[13]. With these 5 biomarkers plus hypoxemia and lymphopenia (1: abnormal; 0: otherwise), we generated a “severity score” ranging from 0 to 7 (values 0 and 1, as well as 6 and 7, were collapsed to assure enough number of patients) which showed a positive linear trend with the hazard ratio of in-hospital mortality (p=0.01), after adjusting for age, sex, baseline characteristics, outpatient treatments, hospital and date of admission (see Supplementary Fig 1).

### In-hospital drug exposure

The main exposure of interest was the inpatient use of RASIs (ACEIs and ARBs), including combinations with other antihypertensive drugs. We also collected information of in-hospital use of the following drugs: calcium channel blockers (CCBs), beta-blocking agents, alpha-adrenoceptor antagonists with cardiovascular (CV) indications, high-ceiling diuretics, low-ceiling diuretics, AMRs, lipid-lowering drugs, glucose-lowering drugs (oral and insulin), antiplatelet drugs, anticoagulants (oral or parenteral), antiviral agents, chloroquine/hydroxychloroquine, azithromycin and other macrolides, other antibiotic agents, systemic steroids and other immunomodulators.

### Outcomes

The main outcome variable was time to in-hospital death for any cause. As a secondary outcome we also considered the time to a composite of in-hospital death and time to ICU admission, whichever occurred first.

### Statistical analysis

We expressed quantitative variables as mean and standard deviation (SD), or median and interquartile range (IQR) for not normally distributed data, and qualitative variables as frequencies and percentages. Differences in quantitative variables were assessed using the Student’s t-test or Mann-Whitney U test (for parametric or non-parametric evaluation between two groups, respectively). Differences in frequencies were assessed using the chi-squared test or Fisher’s exact test when assumptions for chi-square test were not met. The standardized difference was also calculated for means and proportions as a measure of the covariate balance between the exposure groups [21].

To estimate the effect of RASI discontinuation on the outcomes we carried out an intention-to-treat (ITT) analysis, so that patients were analyzed in their assigned closed cohorts (discontinuation or continuation) defined in the first three days of hospitalization, whatever happened thereafter. Then, we proceeded as follows: 1) A binary logistic model was constructed to estimate the propensity score (PS) of RASI discontinuation conditioned on baseline co-morbidities, outpatient treatments, hospital of admission, date of admission (in three periods of equal length), severity score at admission, presence of pneumonia, and treatments prescribed in the first three days of hospitalization (including antihypertensive drugs, chloroquine/hydroxychloroquine, and antivirals, the latter two prescribed per protocol for most admitted COVID-19 patients); 2) Then, we built a Cox proportional hazards model which included the exposure and the estimated PS as a flexible function (restricted cubic splines with 5 knots accounting for 5^th^, 25^th^, 50^th^, 75^th^ and 95^th^ percentiles) to compute the PS-adjusted hazard ratios (HRs) and their 95% confidence intervals (95%CI) [2,23]; 3) We also estimated the controlled direct effect of RASI discontinuation on outcomes by including in the PS-adjusted Cox model the potential mediators (those associated with the exposure, as well as the outcome, controlling for the exposure [23]: systemic corticosteroids, anticoagulants and immunomodulators when death was the outcome and immunomodulators and anticoagulants when the outcome was death plus ICU admission). To avoid a collider bias we also included potential mediator-outcome confounders in the Cox model [24,25] (antiplatelet drugs when the outcome was death and systemic steroids when the outcome was death plus ICU admission), according to our hypothesized causal graph (see Supplementary Fig 2). This way we computed the mediator-controlled HRs (MC-HR) and their 95% CIs.

We also built univariate Kaplan-Meier survival curves for the exposures and outcomes of interest, using log-rank test to evaluate the differences in survival curves across different levels of exposure. The proportional hazard assumption of COX models was checked using the Schoenfeld residuals test and confirmed graphically with a log-minus-log survival plot and by comparison of the Kaplan-Meier survival curves with the Cox predicted curves [23].

The possible effect modification (or interaction) by gender, age, diabetes, obesity, background CV risk, heart failure, severity score (in two categories, using the median as the cut-off point), and in-hospital use of corticosteroids and beta blockers, was assessed stratifying the Cox model by the categories of the potential interacting variables and then comparing the HRs across strata with the Altman and Bland test for interaction [25]. The background CV risk was built as a composite variable with two categories: 1) antecedents of CV disease which includes ischemic heart disease, cerebrovascular accident, heart failure, atrial fibrillation, and thromboembolic disease, and; 2) CV risk factors only which includes hypertension, dyslipidemia, diabetes or chronic renal failure

All the aforementioned analyses were performed for the following comparisons: 1) RASI discontinuation vs RASI continuation; 2) ACEI discontinuation vs. ACEI continuation; 3) ARB discontinuation vs. ARB continuation; and 4) ARB continuation vs. ACEI continuation.

All analyses were performed with STATA/SE v.15 (StataCorp LLC, College Station, TX. USA. 2017) and Python (Python Software Foundation, 2001-2020).

### Sensitivity analyses

Three sensitivity analyses were performed: 1) reclassifying patients in whom RASI discontinuation was uncertain, so that those with a sole prescription recorded in day 2 or day 3 were assigned to the continuation cohort, and patients with a sole prescription recorded in day 1 were assigned to the discontinuation cohort; 2) assigning all patients in whom discontinuation was uncertain to the discontinuation cohort; and 3) using a 2-days window, instead of a 3-days window, to define RASI (dis)continuation (see Supplementary Fig 3).

### Ethics approval

The present study was an extension of a previous study approved by the Ethics Research Committee of the University Hospital “Príncipe de Asturias” on March 18, 2020 (#SRAA-COVID19), including a waiver for the informed consent [6]. The Ethics Research Committee was informed of this extension, and no additional ethical assessment was required. Data extracted were fully anonymised and no attempt was made to interview patients or relatives. The study complied with the provisions of the European and Spanish legislation.

## RESULTS

### Patient selection and discontinuation rates

A total of 2029 patients were consecutively admitted with a PCR-confirmed COVID-19, being 819 outpatient users of RASIs. In 141 of them we were unable to assess the continuation of RASIs (59 patients were directly admitted to the ICU: 47 from the ED and 12 from other hospitals; 44 were transferred from the ED to another hospital; 38 had the event -death or ICU admission- or were discharged within the first 3 days of admission); and in 53 the intention-to-discontinue was uncertain (22 presented a sole prescription in day 1 and 31 in days 2 or 3). Overall, 625 patients were included in the main analysis; out of them, 285 (45.6%) continued and 340 (54.4%) discontinued RASI treatment (Fig 1).

**Figure 1.**
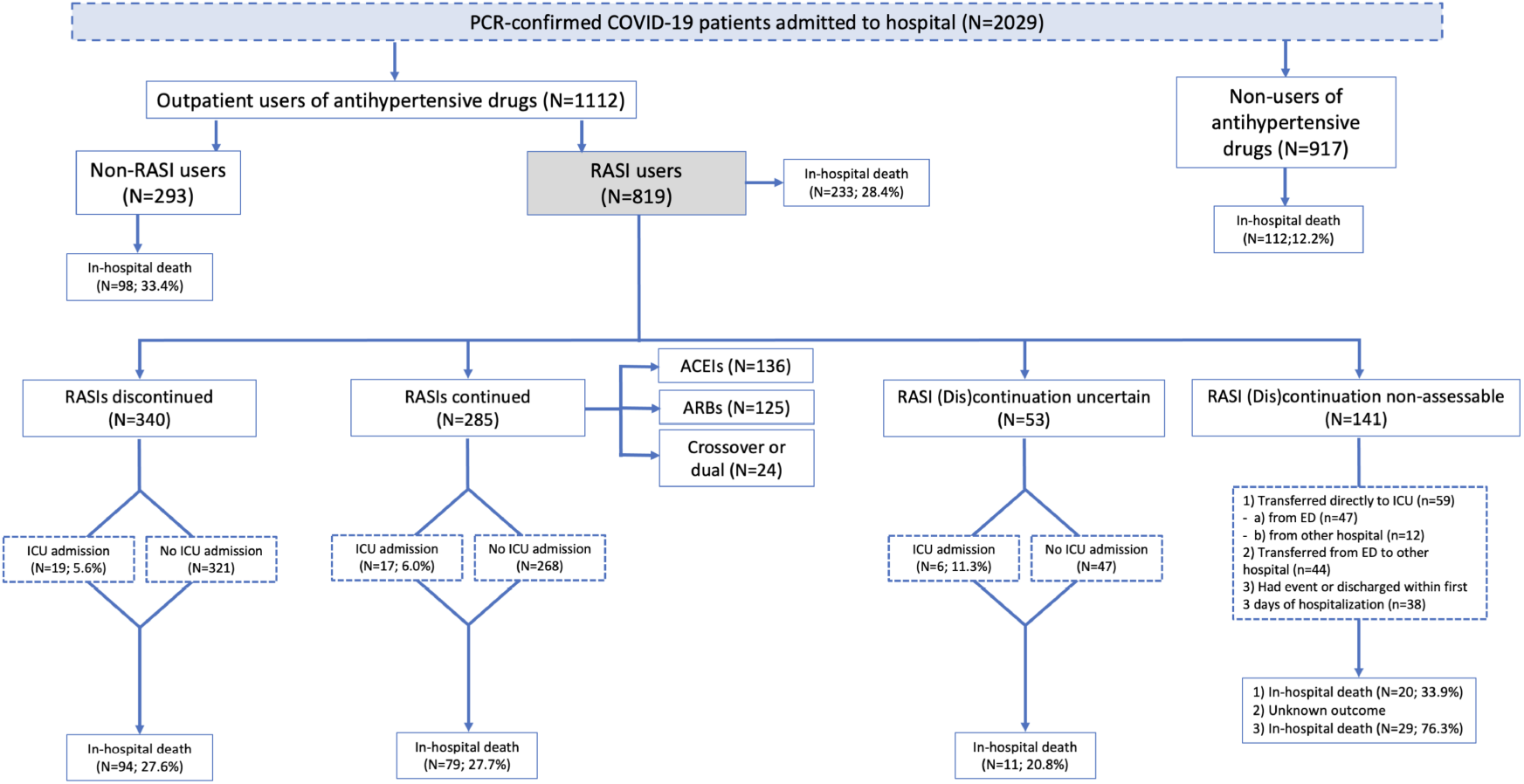
Study design and patient selection. **Abbreviations:** ED: emergency department; ICU: intensive care unit; Non-RASIs: other antihypertensive drugs different from RASIs; RASIs: renin-angiotensin system inhibitors.

RASI discontinuation rates varied greatly across participating hospitals (ranging from 23.5% to 93.0%) and proved to be highly dependent on the date of admission (from 32.1% in the first 10 days of March to 74.2% in last 10 days of March)(Table 1 and Supplementary Fig 4). Among patients who discontinued RASIs, 131 (38.5%) received treatment with CCBs (alone or combined with other antihypertensive drugs), 51 (15.0%) with other antihypertensive drugs (OADs) alone, and 158 (46.5%) had no recorded antihypertensive treatment within the first 3 days of admission (furosemide excluded) (Fig 2). A similar pattern was observed when ACEIs and ARBs were considered separately (Supplementary Fig 5).

**Table 1.** Baseline characteristics and disease severity markers at admission of renin-angiotensin system inhibitors users in discontinuation and continuation cohorts.

|  | **RASIs**<br>**discontinued**<br>**N=340 (%)** | **RASIs**<br>**continued**<br>**N=285 (%)** | **Standardized**<br>**difference** | **p-value** |
| --- | --- | --- | --- | --- |
| Sex, Males | 213 (62.7) | 166 (58.3) | +0.09 | 0.26 |
| Age, years, median (IQR) | 74 (65.5-82) | 75 (68-82) | -0.16 | 0.22 |
| ***Baseline risk factors and comorbidities*** |  |  |  |  |
| Obesity | 57 (16.8) | 86 (30.2) | -0.32 | **<0.001** |
| Smoking: |  |  |  |  |
| Non-smoker | 129 (37.9) | 116 (40.7) |  |  |
| Current smoker | 14 (4.1) | 15 (5.3) | -0.09 | 0.76 |
| Past smoker | 102 (30.0) | 81 (28.4) |  |  |
| Not recorded | 95 (27.9) | 73 (25.6) |  |  |
| Hypertension | 332 (97.6) | 277 (97.2) | +0.03 | 0.72 |
| Diabetes | 123 (36.2) | 111 (38.9) | -0.06 | 0.48 |
| Dyslipidemia | 219 (64.4) | 193 (67.7) | -0.07 | 0.38 |
| Ischemic heart disease | 50 (14.7) | 49 (17.2) | -0.07 | 0.40 |
| Heart failure | 29 (8.5) | 40 (14.0) | -0.17 | **0.03** |
| Atrial fibrillation | 50 (14.7) | 49 (17.2) | -0.07 | 0.40 |
| Thromboembolic disease | 11 (3.2) | 17 (6.0) | -0.13 | 0.10 |
| Cerebrovascular accident | 18 (5.3) | 32 (11.2) | -0.22 | **0.01** |
| COPD | 42 (12.4) | 38 (13.3) | -0.03 | 0.71 |
| Asthma | 27 (7.9) | 20 (7.0) | +0.04 | 0.66 |
| Cancer: |  |  |  |  |
| Antecedents | 32 (9.4) | 30 (10.5) | -0.04 | 0.64 |
| Current | 35 (10.3) | 32 (11.2) | -0.03 | 0.71 |
| Chronic Renal Failure | 38 (11.2) | 34 (11.9) | -0.02 | 0.77 |
| ***Treatment before admission*** |  |  |  |  |
| ACEIs | 172 (50.6) | 149 (52.3) | -0.03 | 0.67 |
| ARBs | 170 (50.0) | 138 (48.4) | +0.03 | 0.69 |
| AMRs | 11 (3.2) | 12 (4.2) | -0.05 | 0.52 |
| CCBs | 115 (33.8) | 81 (28.4) | +0.12 | 0.15 |
| Diuretics | 173 (50.9) | 157 (55.1) | -0.08 | 0.29 |
| Beta-blockers | 81 (23.8) | 73 (25.6) | -0.04 | 0.60 |
| Alpha-blockers | 20 (5.9) | 20 (7.0) | -0.05 | 0.56 |
| Oral anticoagulants | 55 (16.2) | 59 (20.7) | -0.12 | 0.14 |
| Antiplatelet agents | 93 (27.4) | 82 (28.8) | -0.03 | 0.69 |
| NSAIDs | 29 (8.5) | 20 (7.0) | +0.06 | 0.48 |
| Systemic corticosteroids | 25 (7.4) | 10 (3.5) | +0.17 | **0.04** |
| Paracetamol | 177 (52.1) | 173 (60.7) | -0.17 | **0.03** |
| Metamizole | 94 (27.6) | 74 (26.0) | +0.04 | 0.64 |
| Statins | 173 (50.9) | 164 (57.5) | -0.13 | 0.10 |
| Ezetimibe | 14 (4.1) | 12 (4.2) | -0.004 | 0.95 |
| Glucose lowering drugs | 100 (29.4) | 94 (33.0) | -0.08 | 0.34 |
| Insulin | 32 (9.4) | 37 (13.0) | -0.11 | 0.16 |
| **Hospital** (row %): |  |  |  |  |
| HUPA | 136 (64.5) | 75 (35.6) |  |  |
| HUG | 28 (23.5) | 91 (76.5) |  |  |
| HURyC | 56 (67.5) | 27 (32.5) |  |  |
| HCDGU | 28 (41.2) | 40 (58.2) | - | <0.001 |
| HCSC | 30 (46.2) | 35 (53.9) |  |  |
| HULPr | 53 (93.0) | 4 (7.0) |  |  |
| HUPHM | 9 (40.9) | 13 (59.1) |  |  |
| **Date of admission** (row %): |  |  |  |  |
| March, 1-10 | 26 (32.1) | 55 (67.9) |  |  |
| March, 11-20 | 219 (52.6) | 197 (47.4) | - | <0.001 |
| March, 21-31 | 95 (74.2) | 33 (25.8) |  |  |
| **Disease severity at admission** |  |  |  |  |
| Pneumonia | 319 (93.8) | 252 (88.4) | +0.19 | **0.02** |
| Hypoxemia* | 153 (45.0) | 116 (40.7) | +0.09 | 0.28 |
| CRP** | 322 (94.7) | 271 (95.1) | -0.02 | 0.83 |
| Troponin** | 21 (6.2) | 19 (6.7) | -0.02 | 0.80 |
| D-dimer** | 184 (54.1) | 137 (48.1) | +0.12 | 0.13 |
| Procalcitonin** | 140 (41.2) | 79 (27.7) | +0.29 | **<0.001** |
| NT-pro-BNP** | 42 (12.4) | 36 (12.6) | -0.01 | 0.92 |
| Lymphopenia | 192 (56.5) | 162 (56.8) | -0.01 | 0.93 |
| **Severity score\*\*\*:** |  |  |  |  |
| 0-1 | 39 (11.5) | 40 (14.0) |  |  |
| 2 | 66 (19.4) | 74 (26.0) |  |  |
| 3 | 109 (32.1) | 80 (28.1) | - | 0.16 |
| 4 | 80 (23.5) | 66 (23.2) |  |  |
| 5 | 39 (11.5) | 20 (7.0) |  |  |
| 6-7 | 7 (2.1) | 5 (1.8) |  |  |
| Mean (SD) | 3.1 (1.2) | 2.9 (1.2) | +0.18 | **0.03** |

**Abbreviations:** ACEIs: angiotensin-converting enzyme inhibitors; AMRs: antagonists of mineralocorticoid receptor; ARBs: angiotensin receptor blockers; COPD: chronic obstructive pulmonary disease; CCBs: calcium channel blockers; CRP: C-reactive protein; HCDGU: Hospital Central de la Defensa Gómez Ulla; HCSC: Hospital Clínico San Carlos; HUG: Hospital Universitario de Getafe; HULPr: Hospital Universitario La Princesa; HUPA: Hospital Universitario Príncipe de Asturias; HUPHM: Hospital Universitario Puerta de Hierro de Madrid; HURyC: Hospital Universitario Ramón y Cajal; IQR: interquartile range; NSAIDs: nonsteroidal anti-inflammatory drugs; NT-pro-BNP: N terminal proprotein natriuretic peptide type B; RASIs: renin-angiotensin system inhibitors; SD: standard deviation.
\*SatO<sub>2</sub> <90%, or PaO<sub>2</sub>/FiO<sub>2</sub> ≤315 mm Hg
\*\* Patients with abnormal values measured at admission
\*\*\* Composite variable including: hypoxemia, CRP, troponin, D-dimer, procalcitonin, NT-pro-BNP, and lymphopenia. Abnormal value: 1; normal or missing: 0.

**Figure 2:**
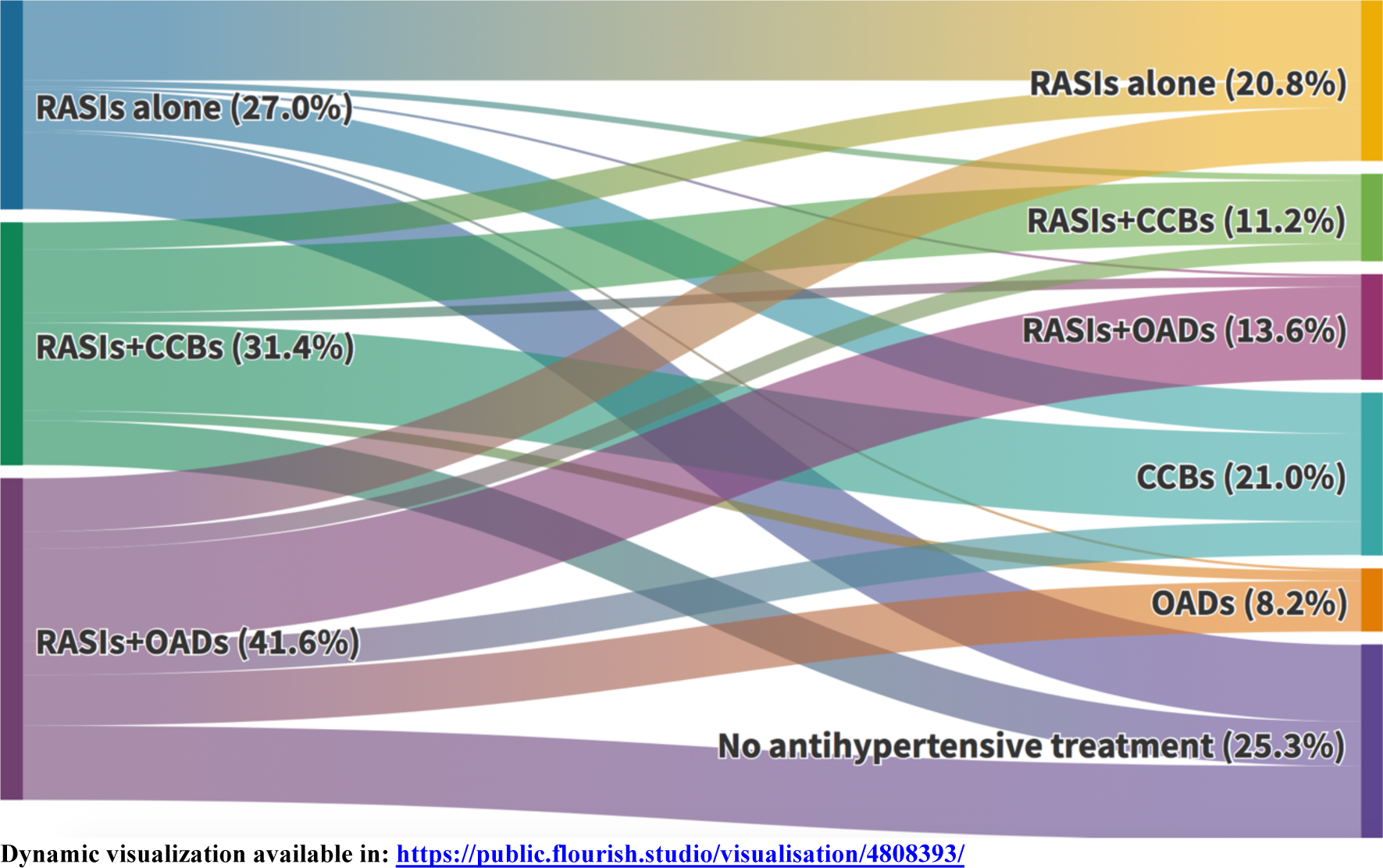
Switching from RASIs to CCBs, other antihypertensive drugs (OADs) or no antihypertensive treatment during the first 3 days since hospital admission (patients with uncertain discontinuation were excluded). Of all outpatient RASI users, 45.6% continued with RASIs (alone or combined with CCBs or OADs), 29.2% were switched to CCBs or OADs, and 25.3% were left without any antihypertensive treatment. **Abbreviations:** CCBs: calcium-channel blockers; OADs: other antihypertensive drugs (different from RASIs or CCBs); RASIs: renin-angiotensin system inhibitors; **RASIs+CCB:** combined use with OADs allowed; **RASIs+OADs:** use of CCBs excluded; **CCBs:** alone or combined with OADs and RASIs excluded; **OADs:** use of RASIs and CCBs excluded.

### Patient characteristics

The baseline characteristics of patients who discontinued and continued treatment with RASIs are shown in Table 1. Baseline co-morbidities and co-medications appeared to be well-balanced, though patients who discontinued had a broadly lower prevalence of co-morbidities (but only statistically significant for obesity, history of heart failure and history of a cerebrovascular accident). At admission, severity markers appeared to be well-balanced, though patients who discontinued presented a slightly higher proportion of pneumonia (93.8% vs. 88.4%; p=0.02), and slightly higher average severity score (3.1 vs. 2.9; p=0.03) (Table 1).

During hospitalization, patients in whom RASIs were discontinued presented a higher proportion of treatment with parenteral anticoagulants, systemic corticosteroids, and CCBs, while patients who continued with RASIs presented a higher use of oral anticoagulants, statins, oral glucose-lowering drugs, other macrolides (different from azithromycin), tocilizumab or other immunomodulating agents, beta-blockers and low-ceiling diuretics (Table 2). ICU admission was similar in both groups (5.6% vs. 6.0% for patients who discontinued and continued with RASIs, respectively), as well as the median hospital stay (11 vs. 10 days). Similar patterns were observed when RASIs were disaggregated by ACEIs and ARBs (Supplementary Tables 1 and 2).

**Table 2.** In-hospital stay and treatment received according to discontinuation or continuation of RASIs.

|  | **RASIs**<br>**discontinued**<br>**n=340 (%)** | **RASIs**<br>**continued**<br>**n=285 (%)** | **Standardized**<br>**difference** | **p-value** |
| --- | --- | --- | --- | --- |
| **Hospital stay, days,**<br>**median (IQR)** | 11 (7.5-17) | 10 (7-16) | +0.01 | 0.17 |
| **ICU admission** | 19 (5.6) | 17 (6.0) | +0.02 | 0.84 |
| **Anticoagulants:** |  |  |  |  |
| Oral | 26 (7.7) | 42 (14.7) | -0.23 | **0.01** |
| Parenteral | 273 (80.3) | 166 (58.2) | +0.49 | **<0.001** |
| **Antiplatelet drugs** | 80 (23.5) | 86 (30.2) | -0.15 | 0.06 |
| **Statins** | 46 (13.5) | 113 (39.7) | -0.62 | **<0.001** |
| **Glucose lowering drugs:** |  |  |  |  |
| Oral | 14 (4.1) | 43 (15.1) | -0.38 | **<0.001** |
| Insulin | 125 (36.8) | 90 (31.6) | +0.11 | 0.17 |
| **Hydroxychloroquine** | 306 (90.0) | 244 (85.6) | +0.13 | 0.09 |
| **Lopinavir+Ritonavir/**<br>**Darunavir+Cobicistat** | 286 (84.1) | 233 (81.8) | +0.06 | 0.43 |
| **Azithromycin** | 128 (37.7) | 116 (40.7) | -0.06 | 0.44 |
| **Other macrolides** | 10 (2.9) | 19 (6.7) | -0.17 | **0.03** |
| **Other antivirals\*** | 8 (2.4) | 6 (2.1) | +0.02 | 0.84 |
| **Other antibacterial agents** | 212 (62.4) | 194 (68.1) | -0.12 | 0.14 |
| **Immunomodulators:** |  |  |  |  |
| Tocilizumab | 43 (12.7) | 57 (20.0) | -0.20 | **0.01** |
| Others** | 99 (29.1) | 121 (42.5) | -0.28 | **<0.001** |
| **Corticosteroids** | 167 (49.1) | 112 (39.3) | +0.20 | **0.01** |
| **Antihypertensive drugs:** |  |  |  |  |
| CCBs | 131 (38.5) | 70 (24.6) | +0.30 | **<0.001** |
| Beta-blockers | 60 (17.7) | 69 (24.2) | -0.16 | **0.04** |
| Alpha-blockers | 11 (3.2) | 9 (3.2) | +0.004 | 0.96 |
| High-ceiling diuretics | 52 (15.3) | 44 (15.4) | -0.004 | 0.96 |
| Low-ceiling diuretics | 17 (5.0) | 53 (18.6) | -0.43 | **<0.001** |

**Abbreviations:** CCBs: calcium channel blockers; ICU: intensive care unit; IQR: interquartile range; RASIs: renin-angiotensin system inhibitors
\* Other antivirals: remdesivir, aciclovir, bicittegravir-emtricitabine-tenofovir, tenofovir, emtricitabine-tenofovir, lamivudine-abacavir-dolutegravir, valaciclovir and valganciclovir.
\*\*Other immunomodulators: Jak inhibitors, interferon beta-1b, ciclosporin, anakinra, ceftriaxone, leflunomide, methotrexate and mycophenolic acid.

### Mortality rates associated with RASI discontinuation vs. RASI continuation

Among patients in whom RASIs were discontinued, 94 (27.6%) died in-hospital whereas 79 (27.7%) died among patients in whom RASIs were continued, which yielded a PS-adjusted HR of 1.01 (95%CI:0.71-1.46) that was not modified after controlling for potential mediators (MC-HR=1.01; 95%CI:0.70-1.46). Similar results were found when the outcome was the composite of death and ICU admission (Table 3). Of note, when RASIs were disaggregated by ACEIs and ARBs, we found a non-significant increased mortality risk among patients in whom ARBs were discontinued (PS-adjusted HR=1.58; 95%CI:0.87-2.87; and MC-HR=1.59; 95%CI:0.89-2.85), whereas among patients in whom ACEIs were discontinued we observed the opposite trend (PS-adjusted HR=0.73; 95%CI:0.44-1.19; and MC-HR=0.70; 95%CI:0.42-1.17) (Table 3).

**Table 3.**
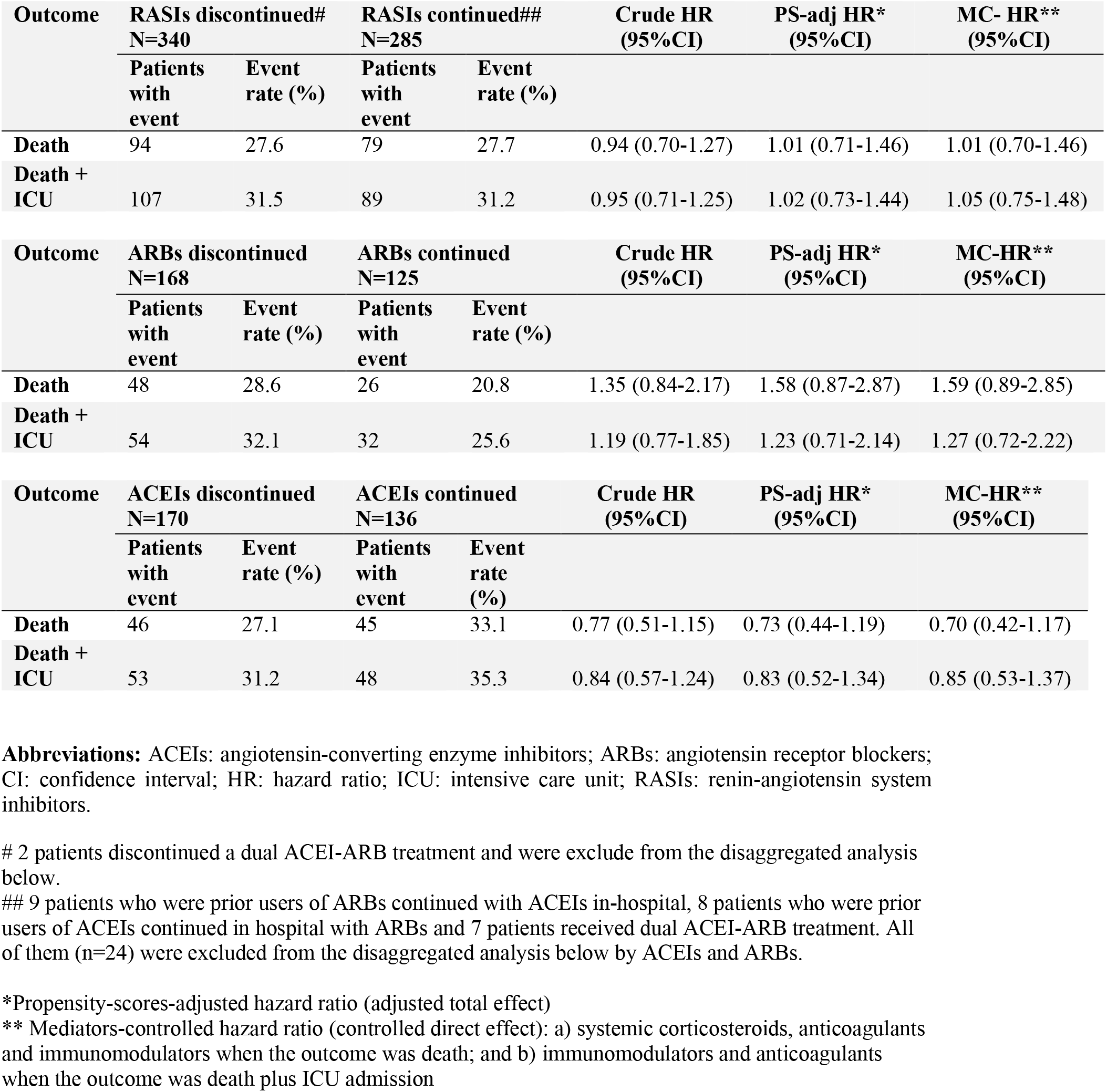
Crude and adjusted hazard ratios of in-hospital death or a composite of death and ICU admission, according to the discontinuation or continuation of in-hospital of ACEIs and ARBs, either pooling as a group (RASI) or disaggregated.

### Head-to-head comparison between ARB versus ACEI continuation

Among 285 patients who continued with RASIs, 136 did so with ACEIs and 125 with ARBs; 24 patients who used dual therapy or were crossed over to the other treatment were excluded from this analysis. The baseline characteristics and in-hospital treatment of patients who continued with ARBs and ACEIs appeared to be evenly distributed (Supplementary Table 3), but the mortality rates were remarkably different (20.8% vs. 33.1% for ARBs and ACEIs, respectively; p=0.03), yielding a head-to-head crude HR of 0.57 (95%CI:0.35-0.93), which barely changed after adjustment for baseline covariates (PS-HR=0.56; 95%CI: 0.32-0.99) and after controlling for mediators (including systemic corticosteroids, immunomodulators and anticoagulants) (MC-HR=0.52; 95%CI:0.29-0.93) (Table 4). The respective Kaplan-Meier survival curves are shown in Fig 3, with the log-rank test resulting in a p value of 0.02. The median survival time was 25 days for patients who continued with ACEIs and was not reached for patients who continued with ARBs. For the composite outcome, the trend to a reduced mortality risk associated with ARBs as compared to ACEIs was still present, but did not reach statistical significance (MC-HR= 0.59; 95%CI:0.35-1.01) (Table 4).

**Table 4.** Head-to-head comparison of patients who continued treatment with angiotensin receptor blockers with patients who continued treatment with angiotensin converting enzyme inhibitors.

| Outcome | ARBs continued<br>N=125 |  | ACEIs continued<br>N=136 |  | Crude HR<br>(95%CI) | PS-adj HR*<br>(95%CI) | MC-HR**<br>(95%CI) |
| --- | --- | --- | --- | --- | --- | --- | --- |
|  | Patients<br>with<br>event | Event<br>rate (%) | Patients<br>with<br>event | Event<br>rate<br>(%) |  |  |  |
| Death | 26 | 20.8 | 45 | 33.1 | 0.57 (0.35-0.93) | 0.56 (0.32-0.99) | 0.52 (0.29-0.93) |
| Death +<br>ICU | 32 | 25.6 | 48 | 35.3 | 0.69 (0.44-1.08) | 0.69 (0.41-1.16) | 0.59 (0.35-1.01) |

**Abbreviations:** ACEIs: angiotensin-converting enzyme inhibitors; ARBs: angiotensin receptor blockers; CI: confidence interval; HR: hazard ratio; ICU: intensive care unit
\*Propensity-scores-adjusted hazard ratio (adjusted total effect)
\*\* Mediators-controlled hazard ratio (controlled direct effect): a) systemic corticosteroids, anticoagulants and immunomodulators when the outcome was death; and b) immunomodulators and anticoagulants when the outcome was death plus ICU admission

**Figure 3.**
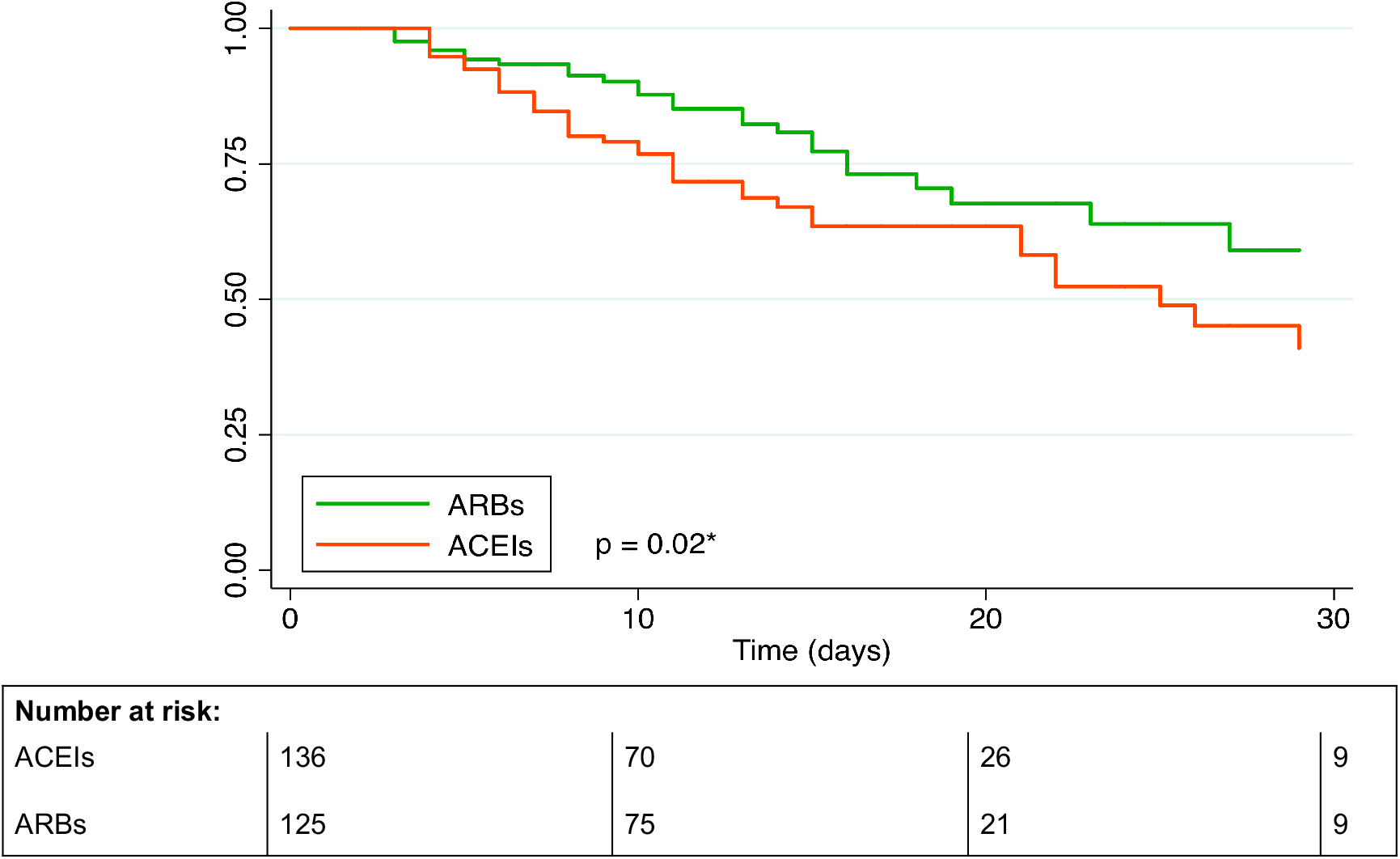
Kaplan-Meier survival curves of in-hospital death among patients in whom treatment with ARBs was continued as compared to those in whom ACEIs was continued (defined in the first 3-days window). **Abbreviations:** ACEIs: angiotensin-converting enzyme inhibitors; ARBs: angiotensin receptor blockers. * Log-rank test

### Analysis of potential interactions

No statistically significant interaction was observed by gender, age (<75; 75+years), obesity, diabetes, heart failure, background cardiovascular risk, severity score (0-3; 4-7) and in-hospital use of corticosteroids or beta-blockers (Supplementary Fig 6). The results disaggregated by ACEIs and ARBs are shown in Supplementary figure 7. A trend to a higher risk associated with ARBs discontinuation was observed in all subgroups, being particularly relevant for obese people (MC-HR= 5.40; 95%CI:1.25-23.3; test for interaction p=0.08)

For the comparison between continuation with ARBs vs continuation with ACEIs, we found a statistically significant interaction with a past history of heart failure (Fig 4). It is interesting to note that the reduced risk of mortality associated with ARBs continuation as compared to ACEI continuation was particularly relevant (and statistically significant) in high-risk subgroups: males, patients aged 75 years or older, obese, diabetics and patients with antecedents of heart failure (Fig 4). It is also important to highlight that the use of in-hospital systemic corticosteroids did not appear to mediate or modify the reduced risk associated with ARBs continuation (MC-HR in patients who received corticosteroids=0.54, 95%CI:0.27-1.09; and MC-HR in patients who did not=0.46 (95%CI:0.17-1.23) (Fig 4).

**Figure 4:**
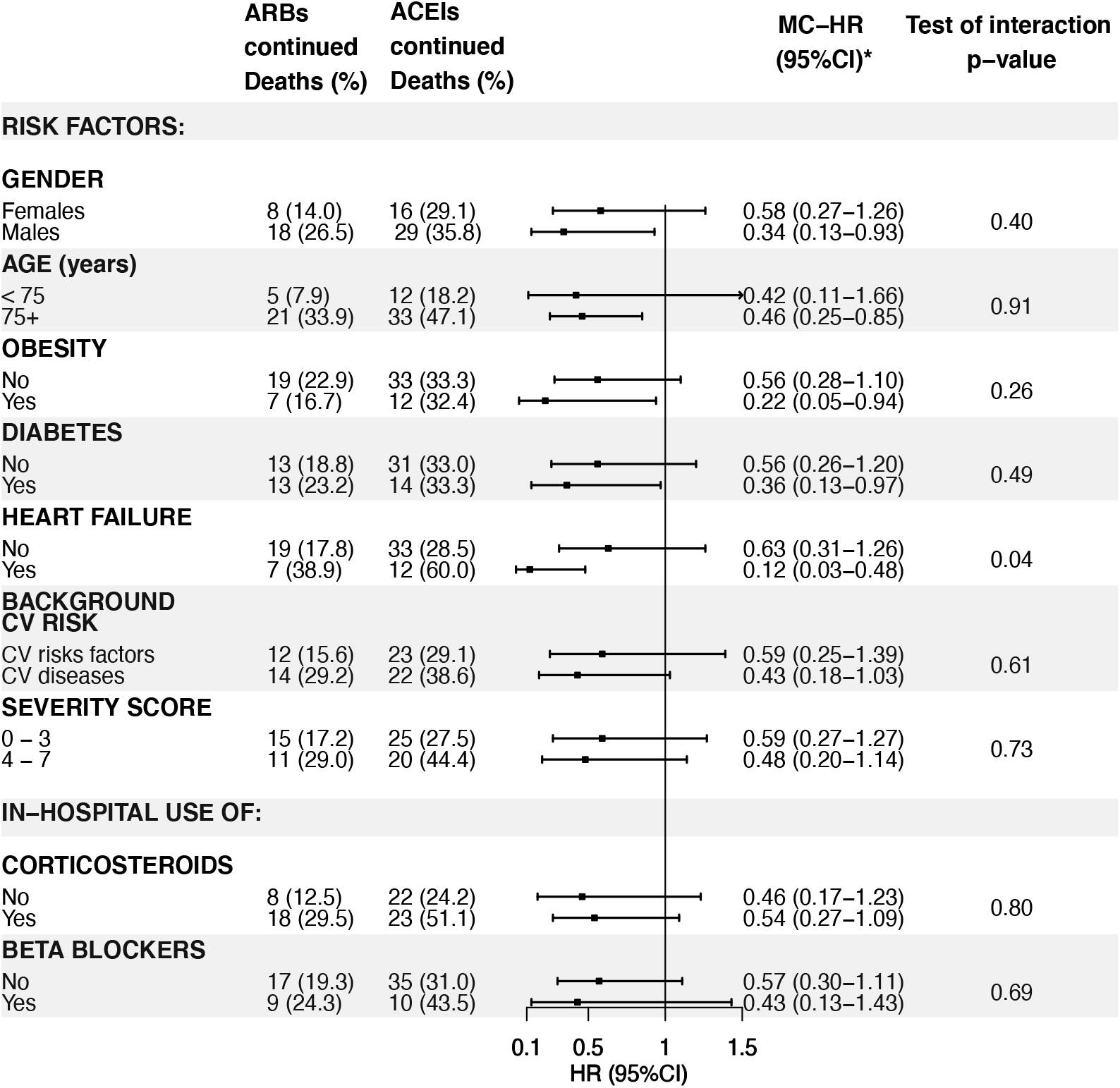
Head-to-head comparison of continuation with angiotensin receptor blockers vs. continuation with angiotensin converting enzyme inhibitors, by different subgroups. **Abbreviations:** ACEIs: angiotensin-converting enzyme inhibitors; ARBs: angiotensin receptor blockers; CV: cardiovascular * Mediators-controlled hazard ratio (controlled direct effect): including systemic corticosteroids (excepting stratification by corticosteroids), anticoagulants and immunomodulators

### Sensitivity analyses

Sensitivity analyses performed after reclassifying patients with uncertain (dis)continuation or using a 2-day window yielded similar results in the main analysis (Supplementary Table 4).

The proportional hazards assumption was fulfilled for all Cox regression analyses according to the Schoenfeld residuals test.

## DISCUSSION

The main findings of the present study are as follows: 1) RASIs were discontinued in around half of patients admitted to hospital for COVID-19 during March, 2020; 2) The discontinuation rate increased over time, being particularly notorious since 11^th^ March; 3) The discontinuation of RASIs as a group was not associated with an increased or decreased risk of in-hospital death or ICU admission, but the results disaggregated by ARBs and ACEIs were not homogeneous; and 4) the continuation of treatment with ARBs was associated with a significantly lower all-cause mortality than the continuation of treatment with ACEIs.

The RASI discontinuation rate was strongly influenced by the date of admission (doubling from mid-March), which seems to be a direct consequence of the hypothesis that quickly spread since 11^th^ March on the possibility that these drugs could make COVID-19 more severe [3]. Notwithstanding, the rate varied considerably by hospital (and possibly by the attending physician within each hospital). In other countries, researchers have reported discontinuation rates ranging from 12.4% to 67.7%, though using different definitions for discontinuation (Supplementary Table 5) [9-12,17, 27-34]. Of note, in our study as much as 46.5% of patients in whom treatment with RASIs were discontinued (25.3% of the total number of patients who used them prior to admission) were left without any antihypertensive drug (excluding furosemide), which suggests that in a relevant part of patients RASIs were discontinued for medical reasons, likely related to an unstable hemodynamic situation.

Our main finding is that the discontinuation of RASIs, as a group, did not have an impact on in-hospital mortality or in the composite of in-hospital mortality plus ICU admission. This result seems robust as it hardly varied in different sensitivity analyses in which we modified the definition of (dis)continuation. Contrary to the huge number of studies carried out to assess the impact of outpatient use of RASIs on different outcomes (COVID-19 infection, hospitalization and mortality, among others) [7,8], fewer studies have been performed thus far to examine the association of inpatient use of RASIs with in-hospital mortality. One of the first studies was published by Zhang et al. [11] with data from 9 hospitals in Hubei province (China). They found an all-cause mortality among inpatients treated with RASIs much lower than non-treated patients, with an adjusted HR of 0.42 (95%CI: 0.19-0.92). However, this study was criticized because the authors considered exposed to all patients who received RASIs at any time point during hospitalization, which implies that exposed patients had to survive long enough, or be clinically stable enough, to receive the treatment with RASIs [16]. Thus, such definition of the exposure could have introduced an immortal-time bias [16] and a confounding by severity (also graphically called “healthy user-sick stopper” bias” [17], that is, RASIs were more likely to be continued, initiated or reinstated in less severe cases), both favoring an overestimation of the benefit of RASIs on mortality. Most researchers thereafter used similar definitions incurring in the same types of bias and most coinciding to show an important reduced mortality risk associated with RASIs [9,10,12,27-34] (see supplementary Table 5 for a detailed description of studies). To overcome these problems, we defined continuation or discontinuation during the first 3 days (or during the first 2 days in a sensitivity analysis) and then followed an ITT analysis (each patient analyzed in his/her assigned closed cohort), as it would have been done in a clinical trial. Also, to avoid a reverse causation, we excluded patients directly admitted to the ICU (from the ED or from another hospital), situation in which RASIs are usually discontinued as a consequence of the disease severity. Interestingly, if we had defined continuation as “use of RASIs at any time point during hospitalization” and included patients directly admitted to the ICU in the discontinuation cohort, the mortality rates would have been 25.3% and 30.3% in the continuation and discontinuation cohorts, respectively, yielding a HR of 0.83 (95%CI: 0.66-1.05) for in-hospital mortality. For the composite outcome (death plus ICU admission) the rates would have been 30.0% for patients in whom RASIs were continued and 43.6% in those who discontinued giving rise to a HR of 0.67 (95%CI: 0.57-0.83). Therefore, the results would have been dramatically different than the ones we actually obtained, showing the extent of such biases.

Recently, the results from two randomized clinical trials in which regular users of RASIs who were admitted to hospital for COVID-19 were assigned to discontinuation or continuation arms, have been reported (BRACE-CORONA [18] and REPLACE COVID [19] trials) and both found no difference in the mortality rates, supporting our results.

However, it is important to emphasize that in the BRACE-CORONA trial the mortality rates were very low (2.7% among patients assigned to discontinuation and 2.8% in those assigned to continuation), casting doubts on the generalizability of their results (the mean age of the study population was 55 years old, 20 years younger than our population). Also, the measure of association of mortality was too imprecise (odds ratio=0.97; 95% CI, 0.38-2.52) to be informative. Interestingly, 80% of patients were prior users of ARBs, and the authors found quasi-significant results favoring continuation in older persons, obese patients and in those clinically more severe. The REPLACE COVID trial had a more representative population and consistently found no difference in all-cause mortality (15% and 13% in the continuation and discontinuation arms, respectively). Unfortunately, the sample size was too small to make a meaningful separate analysis by ACEIs and ARBs.

The different mortality rates among patients who continued with ACEIs versus those who continued with ARBs is a novel finding that merits specific comments. Firstly, it is important to emphasize that this comparison is ideal for several reasons: a) these drugs have overlapping indications, thereby the subjects who use them are highly comparable, seemingly reducing by design the possibility of confounding (due to either known and unknown factors); b) the possibility of an immortal-time bias is inexistent, as the same definition of continuation was applied to both cohorts; c) the possibility of a confounding by severity is unlikely, as it is not reasonable to think that physicians used different criteria for the continuation of ARBs or ACEIs, and additionally we applied an ITT analysis once continuation was defined based on the records of the first 3 days of hospitalization; and, finally, d) the few differences we found (such as the greater in-hospital use of systemic corticosteroids in the ARB continuation cohort) were controlled for by including this factor in the outcome regression model and by stratification, and none of these strategies changed the results, reinforcing the internal validity of the comparison.

Secondly, most previous studies have pooled ACEIs and ARBs (see supplementary Table 5), as if they were the same type of drugs. However, our results show that this approach may be wrong; also, there are profound pharmacological reasons that make this grouping invalid, in particular for COVID-19 patients. ARBs block selectively the action of angiotensin II on AT1 receptor (AT1R) and free angiotensin II is then converted by the ACE2 into angiotensin (1-7) which would act on Mas1 receptor (Mas1R) to induce opposite actions to angiotensin II (anti-inflammatory, anti-oxidant, anti-fibrotic, anti-thrombotic, anti-hypertrophic, vasodilatation and natriuresis) [13-15]. Also, angiotensin II not used in activating AT1R, acts on AT2 receptor (AT2R) (for which ARBs have no affinity), whose activation is known to produce opposite actions to the ones derived from the activation of AT1R [15], thereby collaborating with the protective effect of angiotensin (1-7). Instead, ACEIs inhibit the formation of angiotensin II, which preempts the generation of angiotensin (1-7) from both angiotensin II via ACE2, but also from angiotensin (1-9) via ACE1 [13-15]; additionally, the beneficial actions derived from activation of AT2R do not take place. In sum, both ARBs and ACEIs effectively block RAS, whereas only ARBs appear to reinforce its counterpart, via ACE2-angiotensin(1-7)-Mas1R axis and AT2R activation, a difference that could be critical in COVID-19 patients. Additionally, ACE1 is well-known to be the major vascular peptidase of bradykinin, an abundant peptide which promotes vasodilatation, vascular permeability and liberation of inflammatory cytokines (IL-1, IL-2, IL-6. IL-8 and TNF-alpha) implicated in the cytokine storm associated with the severe forms of COVID-19 [15]. Therefore, ACEIs will reduce bradykinin degradation, thereby potentiating its effects, which ultimately could be detrimental for COVID-19 patients [15,35,36]. These negative collateral actions of ACEIs may offset the benefits derived from the inhibition of angiotensin II formation and, we postulate, that these could account for the important difference we found in the mortality rates among inpatients treated with ARBs and those treated with ACEIs (an absolute difference of 12.3%, corresponding to a number needed to treat as low as 8). Importantly, the benefit of ARBs seems to be particular evident in high-risk subgroups: males, the very old, obese, diabetics and patients with antecedents of heart failure. Nevertheless, our results need confirmation, in particular through randomized clinical trials. Some are in progress aiming to assess the benefits of using ARBs in COVID-19 patients with acute respiratory syndrome as compared to placebo [CLARITY (NCT04394117) and NCT04312009], but, as far as we know, no study has been designed to compare ARBs with ACEIs in this context. Rodilla et al [30] compared survival of COVID-19 patients according to the use of ARBs and ACEIs prior to admission and found a significant reduced mortality risk with the former (25.6% vs. 30.4%, respectively, p=0.0001); but, unfortunately, a head-to-head comparison of in-hospital use of ARBs vs. ACEIs was not reported. Finally, it is of interest to note that in the study by Zhang et al[11], 83.5% of patients reported to be on RASIs were actually treated with ARBs.

Our study has some limitations that must be discussed: 1) as in all observational studies, the possibility exists that there is some residual confounding due to unknown or unmeasured factors. Notwithstanding, it is important to remark that all our patients were users of RASIs prior to admission and were highly comparable at baseline, as shown by the good balance of covariates and the fact that the mean and median of the propensity scores for RASI discontinuation was close to 0.5 (not shown); indirectly, it is likely that unmeasured confounding variables are evenly distributed too, albeit this cannot be assured; as previously commented, this is specially applicable to the comparison of ACEI and ARB continuation cohorts; 2) the information on some severity biomarkers (i.e. interleukins 6 or 1β) were not routinely performed at that time and were not considered in the severity score built for this study; on this regard, we would like to emphasize that such score was created to reduce the number of covariates included in the PS models, and it is not proposed as a prognostic index (as we are quite aware that a specific and independent validation study would be necessary for that); 3) the study period selected (March, 2020) was the most critical of the first wave in Spain and, at that time, health professionals worked under an extraordinary pressure, which may have led to under-recording of some relevant clinical information; this limitation, however, does not apply to drugs as they were prescribed through an electronical tool, making unlikely the misclassification of drug exposure; and 4) the mortality rates recorded in our study were extraordinary high (partly accounted for the lack of preparedness of the health system to address this disease at the very beginning of the pandemic) and remarkably different from figures corresponding to other periods during the first and successive waves in Spain or in other countries, so the generalizability of our data on this regard cannot be assured; however, we do not think that this affects the internal validity of our results.

## CONCLUSIONS

The discontinuation of RASIs at hospital admission was common place in the first wave of COVID-19 pandemic in Spain, influenced by the widely spread hypothesis that postulated a more severe disease in patients treated with these drugs. Our results show that the continuation of these drugs during hospitalization did not increase the risk of in-hospital death. On the contrary, we found that the continuation of treatment with ARBs was associated with a trend to a reduced mortality risk as compared to their discontinuation and a significantly lower mortality risk as compared to the continuation with ACEIs, particularly evident in high-risk subgroups. Though further studies are needed, this finding strongly supports the continued use of ARBs in prior users of these drugs admitted for COVID-19, unless medically contraindicated. In regular users of ACEIs, the possibility of switching to ARBs, if admitted to hospital for COVID-19, should be considered, unless medically contraindicated.

## Data Availability

Data sharing: After publication, the data will be made available to others on reasonable requests to the corresponding author. A proposal with detailed description of study objectives and statistical analysis plan will be needed for evaluation of the reasonability of requests. Additional materials might also be required during the process of evaluation. Deidentified participant data will be provided after approval from the principal researchers of the participating hospitals.

## ABBEVIATIONS

ACE1: Angiotensin converting enzyme 1
ACE2: Angiotensin converting enzyme 2
ACEIs: Angiotensin-converting enzyme inhibitors
AMRs: Antagonists of mineralocorticoid receptor
ARBs: Angiotensin receptor blockers
AT1R: Receptor for angiotensin II type 1
AT2R: Receptor for angiotensin II type 2
BMI: Body mass index
CCBs: Calcium channel blockers
CI: Confidence interval
COPD: Chronic obstructive pulmonary disease
COVID-19: Coronavirus disease 19
CPR: C-protein reactive
CV: Cardiovascular
ED: Emergency Department
HR: Hazard ratio
ICU: Intensive care unit
IQR: Interquartile range
ITT: Intention-to-treat
MC-HR: Mediator-controlled hazard ratio
NSAIDs: Nonsteroidal anti-inflammatory drugs
NT-pro-BNP: N terminal pro-peptide of natriuretic factor type B
OADs: Other antihypertensive drugs different from RASIs or CCBs
OR: Odds ratio
PS: Propensity scores
PS-adjusted HR: Propensity score adjusted hazard ratio
RASIs: Renin-angiotensin system inhibitors
SD: Standard deviation

## Acknowledgments

The authors would like to thank health professionals, patients and administrative staff from hospitals participating in this study.

## Disclaimer

The results, discussion and conclusions are from the authors and do not necessarily represent the position of their Institutions.

## Data sharing

After publication, the data will be made available to others on reasonable requests to the corresponding author. A proposal with detailed description of study objectives and statistical analysis plan will be needed for evaluation of the reasonability of requests. Additional materials might also be required during the process of evaluation. Deidentified participant data will be provided after approval from the principal researchers of the participating hospitals.

## AUTHORS’ CONTRIBUTIONS

**Conceptualization and design**: Francisco J. de Abajo

**Data retrieval:** All members of the MED-ACE2-COVID19 Study Group

**Data curation:** Sara Rodríguez-Martín, Antonio Rodríguez-Miguel,

**Formal analysis**: Antonio Rodríguez-Miguel, Sara Rodríguez-Martín and Francisco J de Abajo

**Funding adquisition**: Francisco J. de Abajo

**Investigation:** All members of the MED-ACE2-COVID19 Study Group

**Methodology**: Francisco J. de Abajo, Antonio Rodríguez-Miguel

**Project administration**: Francisco J. de Abajo, Sara Rodríguez-Martín

**Supervision:** Francisco J. de Abajo, Sara Rodríguez-Martín

**Visualization:** Antonio Rodríguez-Miguel, Sara Rodríguez-Martín, Francisco J. de Abajo

**Writing – original draft:** Francisco J. de Abajo, Antonio Rodríguez-Miguel

**Writing -review and editing**: Francisco J. de Abajo, Antonio Rodríguez-Miguel, Sara Rodríguez-Martín, Victoria Lerma and Alberto García-Lledó.

**Final approval for submission:** All members of the MED-ACE2-COVID19 Study Group

