## Supplemental material for "IN-HOSPITAL CONTINUATION WITH ANGIOTENSIN RECEPTOR BLOCKERS IS ASSOCIATED WITH A LOWER MORTALITY RATE THAN CONTINUATION WITH ANGIOTENSIN CONVERTING ENZYME INHIBITORS IN COVID-19 PATIENTS: A RETROSPECTIVE COHORT STUDY"

Prof Francisco J. de Abajo, MD, PhD, MPH[1,2], Antonio Rodríguez-Miguel, PhD[1,2], Sara Rodríguez-Martín, PhD[1,2], Victoria Lerma, RN[1], Alberto García-Lledó, MD, PhD[3,4], on behalf of MED-ACE2-COVID19 Study Group*

1. Clinical Pharmacology Unit. University Hospital Príncipe de Asturias. Alcalá de Henares. Madrid. Spain
2. Department of Biomedical Sciences (Pharmacology Section). University of Alcalá (IRYCIS). Alcalá de Henares. Madrid. Spain
3. Department of Cardiology, University Hospital Príncipe de Asturias. Alcalá de Henares. Madrid. Spain
4. Department of Medicine, University of Alcalá (IRYCIS). Alcalá de Henares. Madrid. Spain.

**Short title:** In-hospital use of RAS inhibitors and mortality in COVID-19 patients

**Key words**: Renin-angiotensin system inhibitors, angiotensin-converting enzyme inhibitors, angiotensin receptor blockers, COVID-19, mortality, in-hospital treatment

**Author for correspondence:**
Prof. Francisco J. de Abajo
Clinical Pharmacology Unit
University Hospital Principe de Asturias
Department of Biomedical Sciences, University of Alcalá (IRYCIS)


***MED-ACE2-COVID19 Study Group:**
*Hospital Universitario Príncipe de Asturias*: F J de Abajo, A Rodríguez-Miguel, S Rodríguez-Martín, V Lerma, A García-Lledó, D Barreira-Hernández; D Rodríguez-Puyol; *Hospital Universitario de Getafe*: O Laosa, L Pedraza, L Rodríguez-Mañas; *Hospital Universitario Ramón y Cajal:* M Aguilar, I de Pablo, MA Gálvez; *Hospital Central de la Defensa Gómez Ulla*: A García-Luque, M Puerro; RM Aparicio, V García-Rosado, C Gutiérrez-Ortega; *Hospital Clínico San Carlos*: L Laredo, E González-Rojano, C Pérez, A Ascaso, C Elvira; *Hospital Universitario de La Princesa*: G Mejía-Abril, P Zubiaur, E Santos-Molina, E Pintos-Sánchez, M Navares-Gómez; F Abad-Santos; *Hospital Universitario Puerta de Hierro-Majadahonda*: G A Centeno, A Sancho-Lopez, C Payares-Herrera, E Diago-Sempere.

**Figure S1. Relationship between the severity score and in-hospital mortality**. Adjusted-hazard ratios of severity score and in-hospital death were obtained trough a regression Cox model after adjusting for age, sex, baseline characteristics, outpatient treatments, hospital and date of admission. Scores 0 and 1, as well as 6 and 7 were collapsed to assure enough number of patients. Test for trend, p=0.01

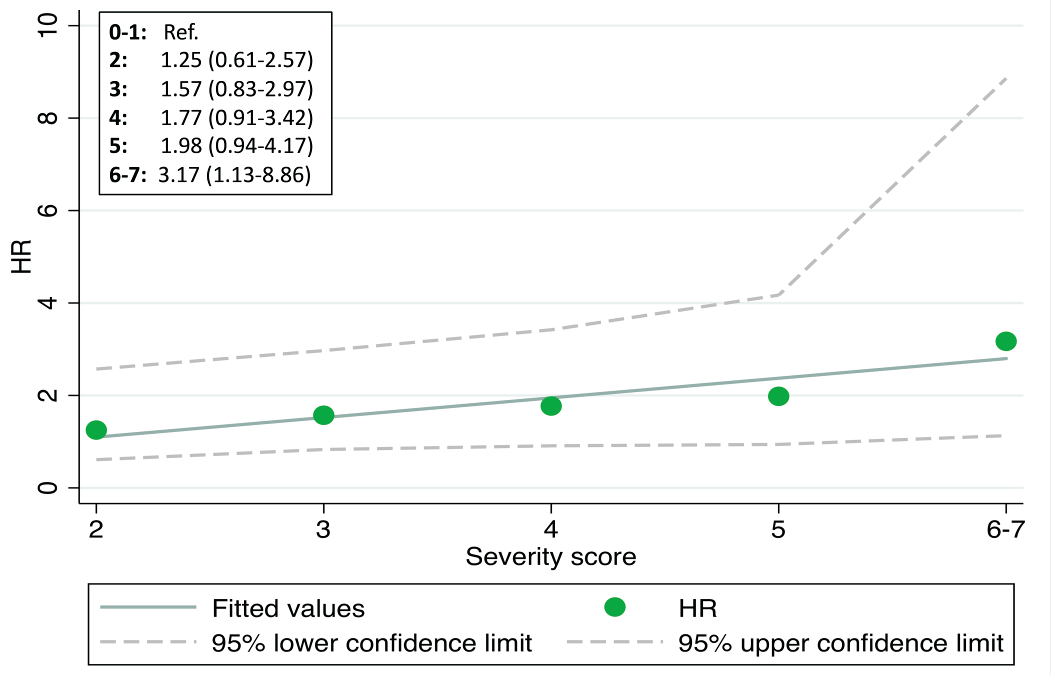

**Figure S2**: **Hypothesized causal graphs.** X: Discontinuation of RASIs; P: Propensity scores; Z: mediators; Y: outcome; and U: mediator-outcome confounder. The arrow connecting X with Y represents the direct effect of X on Y. The pathway through Z is the indirect effect of X on Y (dashed line). In **a)** it is shown the causal diagram when the estimated effect of X on Y is conditioned (indicated by the square) on the propensity scores (adjustment at once for the set of covariates included in the PS model), but no control for Z or adjustment for U is made; so, we would be estimating the total effect of X on Y, including the effect mediated by Z (additionally confounded by U); in **b)** the estimated effect of X on Y is conditioned on P and Z (controlled by the mediator), but the collider on Z opens a path through U and introduces confounding; in **c)** the effect of X on Y is conditioned on P, Z and U, and the direct effect is estimated by controlling for the mediator and adjusting for the mediator-outcome confounder.

**a)**

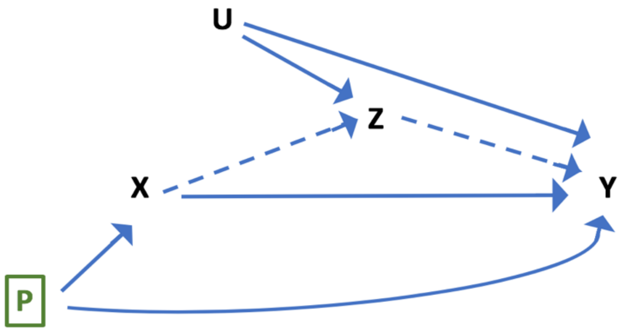


**b)**

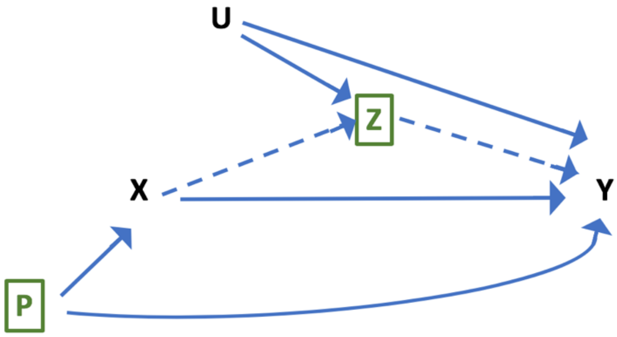


**c)**

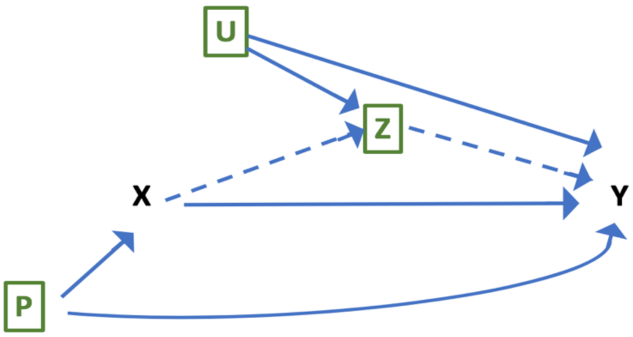

**Figure S3. Definition of discontinuation/continuation of RASIs using: a)** using a 3-days window (main analysis); **b)** using a 2-days window (sensitivity analysis)

**a)**

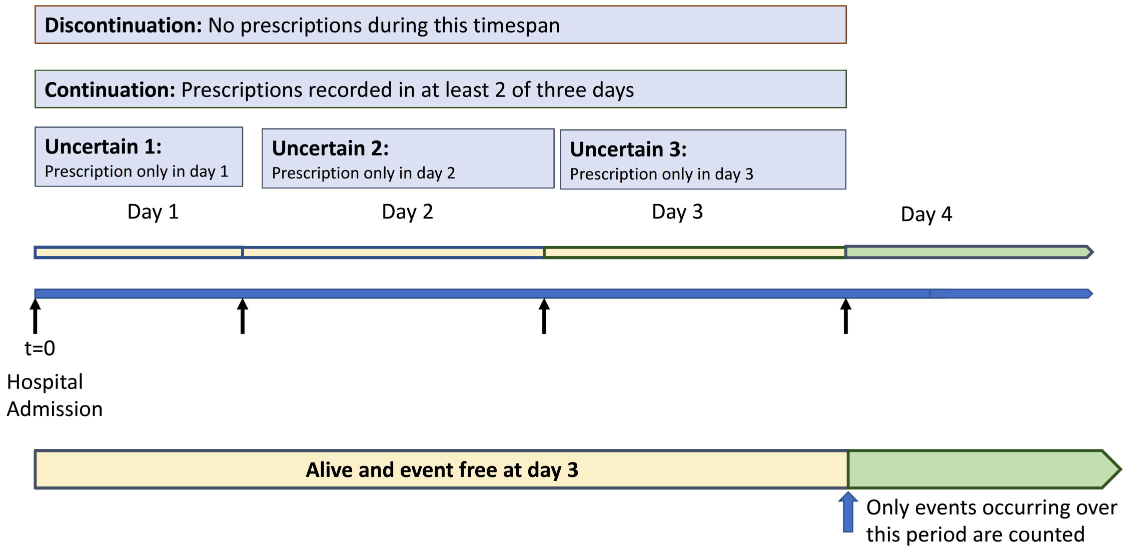


**b)**

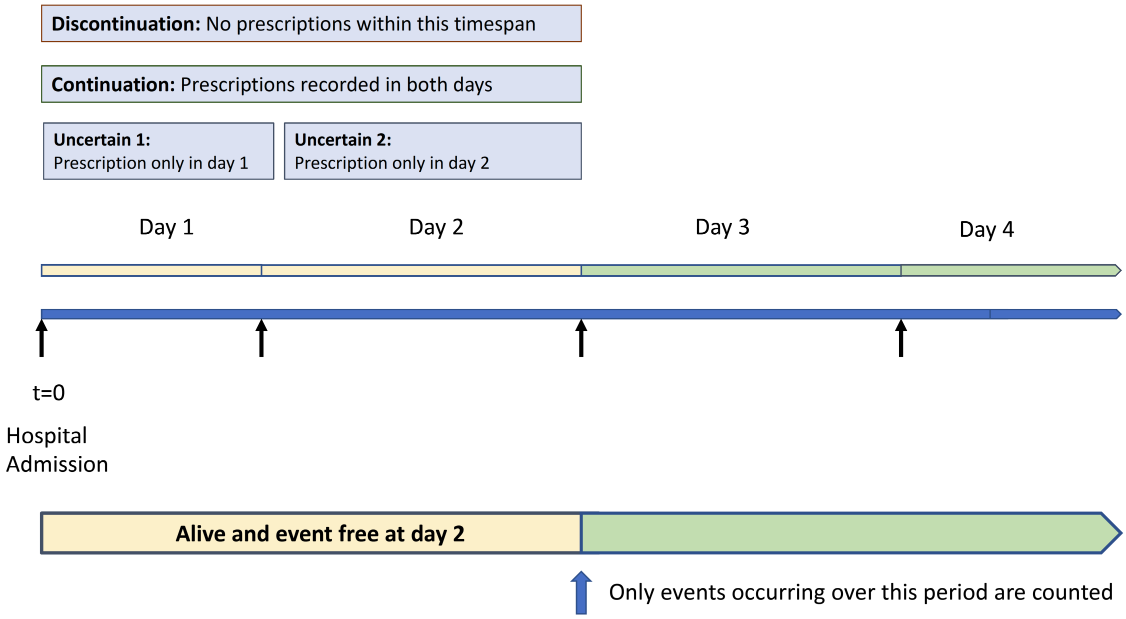

**Figure S4: In-hospital use of RASIs by date-of-admission cohort.** Patients were grouped in three cohorts according to the date of admission: P1) from 1st to 10th of March (n=81); P2) from 11th to 20th of March (N=416); and P3) from 21st to 31st of March (N=128). The discontinuation rate of RASIs was highly influenced by date of admission. The inpatient use of RASIs remained low during hospital stay in all three cohorts.

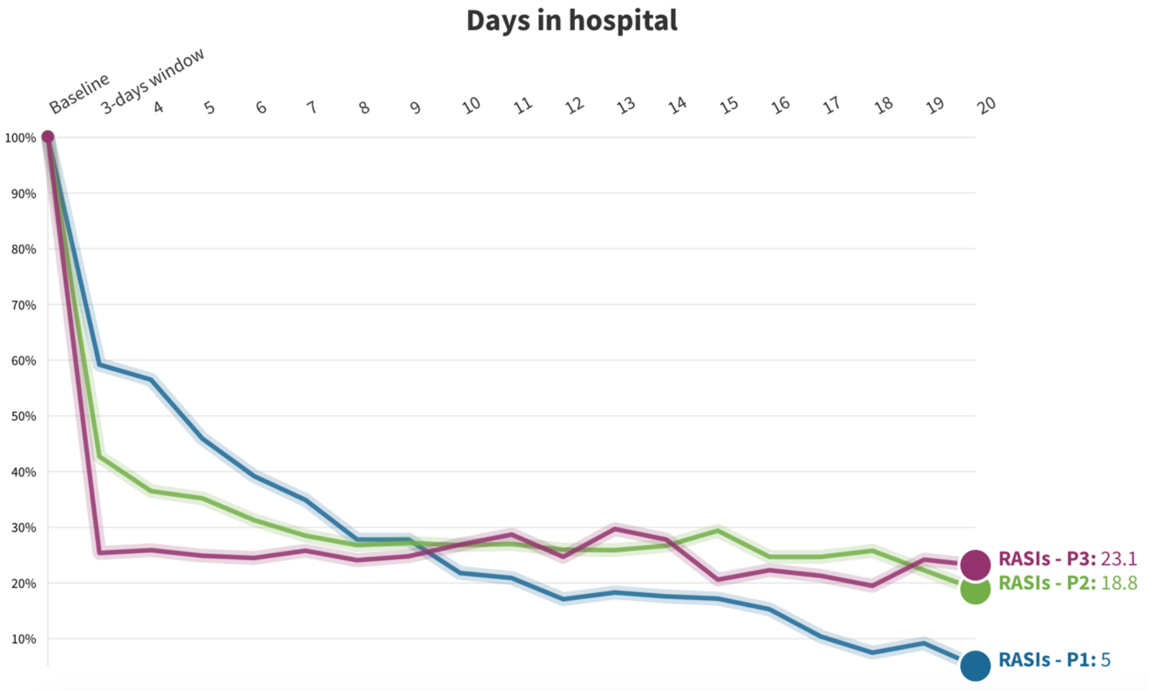


**Dynamic visualization available in:** https://public.flourish.studio/visualisation/4863807/

**Abbreviations:** RASIs: renin-angiotensin system inhibitors

**Figure S5: Switching from ACEIs or ARBs to CCBs and other antihypertensive drugs during the first 3 days since hospital admission (patients with uncertain (dis)continuation are excluded).** Of all outpatient ARB users, 42.7% continued with ARBs (alone or combined with CCBs or OADs), 31.7% were switched to CCBs or OADs, and 25.6% were left without any antihypertensive treatment. Of all outpatient ACEI users, 44.5% continued with ACEIs (alone or combined with CCBs or OADs), 28.8% were switched to CCBs or OADs, and 26.8% were left without any antihypertensive treatment.

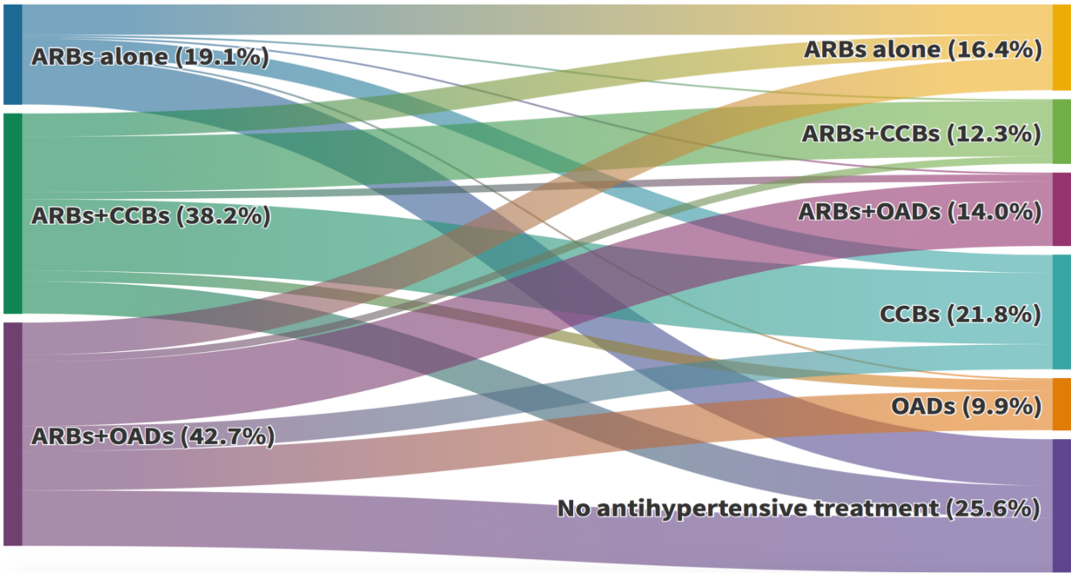


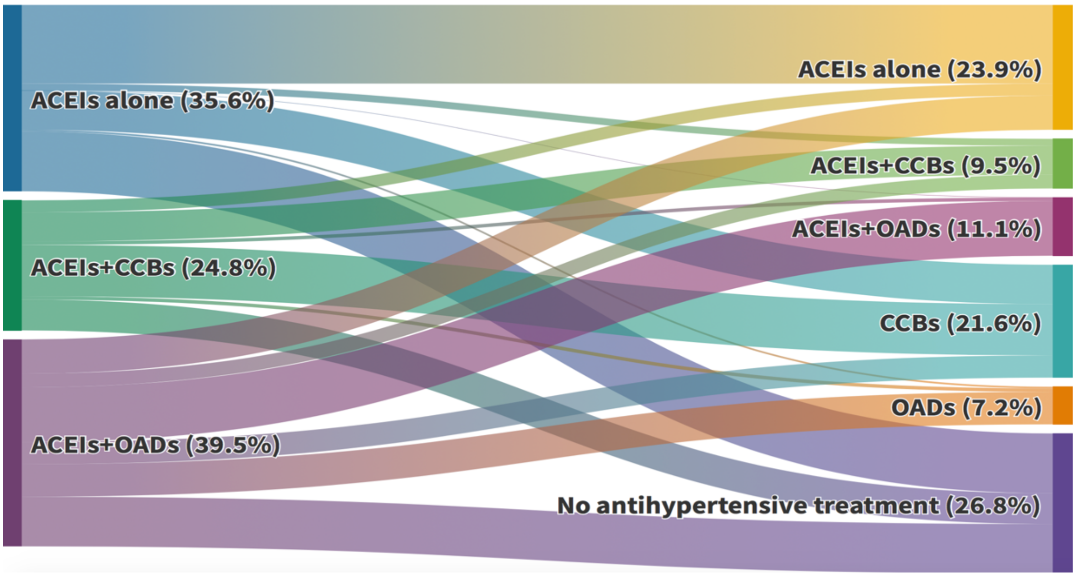


**Dynamic visualizations available in:** ARBs: https://public.flourish.studio/visualisation/4886927/
ACEIs: https://public.flourish.studio/visualisation/4887174/

**Abbreviations:** CCBs: calcium-channel blockers; OADs: other antihypertensive drugs (different from ARBs/ACEIs or CCBs); ARBs: angiotensin receptor blockers; ACEIs: angiotensin-converting enzyme inhibitors

**ARBs/ACEIs+CCB:** combined use with OADs allowed; **ARBs/ACEIs+OADs:** use of CCBs excluded; **CCBs:** alone or combined with OADs and ARBs/ACEIs excluded; **OADs:** use of ARBs/ACEIs and CCBs excluded.

**Figure S6. Forest plots showing mortality rates and adjusted hazard ratios of in-hospital death associated with renin-angiotensin system inhibitors discontinuation vs continuation, by different subgroups.**

| | RASIs discontinued Deaths (%) | RASIs continued Deaths (%) | MC–HR (95%CI)* | Test of interaction p–value |
|---|---|---|---|---|
| **RISK FACTORS:** | | | | |
| **GENDER** | | | | |
| Females | 36 (28.4) | 28 (23.5) | 0.96 (0.53–1.76) | 0.73 |
| Males | 58 (27.2) | 51 (30.7) | 1.10 (0.69–1.76) | |
| **AGE (years)** | | | | |
| < 75 | 21 (12.3) | 19 (13.8) | 1.25 (0.59–2.66) | 0.50 |
| 75+ | 73 (43.2) | 60 (40.8) | 0.93 (0.61–1.44) | |
| **OBESITY** | | | | |
| No | 74 (26.2) | 57 (28.6) | 0.91 (0.60–1.38) | 0.24 |
| Yes | 20 (35.1) | 22 (25.6) | 1.60 (0.74–3.44) | |
| **DIABETES** | | | | |
| No | 55 (25.4) | 46 (26.4) | 0.92 (0.58–1.47) | 0.69 |
| Yes | 39 (31.7) | 33 (29.7) | 1.07 (0.60–1.92) | |
| **HEART FAILURE** | | | | |
| No | 80 (25.7) | 58 (23.7) | 1.07 (0.70–1.61) | 0.38 |
| Yes | 14 (48.3) | 21 (52.5) | 0.71 (0.31–1.59) | |
| **BACKGROUND CV RISK** | | | | |
| CV risks factors | 49 (21.7) | 40 (23.8) | 0.85 (0.51–1.41) | 0.33 |
| CV diseases | 45 (39.5) | 39 (33.3) | 1.22 (0.72–2.07) | |
| **SEVERITY SCORE** | | | | |
| 0 – 3 | 50 (23.4) | 43 (22.2) | 1.06 (0.64–1.77) | 0.88 |
| 4 – 7 | 44 (34.9) | 36 (39.6) | 1.00 (0.58–1.73) | |
| **IN–HOSPITAL USE OF:** | | | | |
| **CORTICOSTEROIDS** | | | | |
| No | 43 (24.9) | 36 (20.8) | 1.19 (0.69–2.06) | 0.32 |
| Yes | 51 (30.5) | 43 (38.4) | 0.82 (0.50–1.35) | |
| **BETA BLOCKERS** | | | | |
| No | 72 (25.7) | 56 (25.9) | 0.87 (0.57–1.33) | 0.39 |
| Yes | 22 (36.7) | 23 (33.3) | 1.26 (0.61–2.59) | |

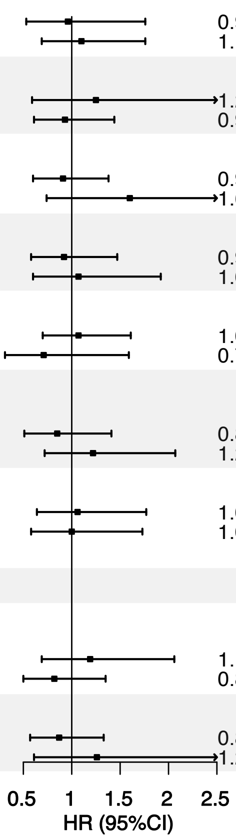


**Abbreviations:** CV: cardiovascular; RASIs: renin-angiotensin system inhibitors

* Mediators-controlled hazard ratio (controlled direct effect): including systemic corticosteroids (excepting stratification by corticosteroids), anticoagulants and immunomodulators

**Figure S7: Forest plots showing mortality rates and adjusted hazard ratios of in-hospital death associated with ACEIs discontinuation vs continuation and ARBs discontinuation vs continuation, by different subgroups.**

| | ACEIs discontinued Deaths (%) | ACEIs continued Deaths (%) | MC–HR (95%CI)* | Test of interaction p–value |
|---|---|---|---|---|
| **RISK FACTORS:** | | | | |
| **GENDER** | | | | |
| Females | 19 (32.2) | 16 (29.1) | 0.85 (0.43–1.68) | 0.56 |
| Males | 27 (24.3) | 29 (35.8) | 0.62 (0.28–1.38) | |
| **AGE (years)** | | | | |
| < 75 | 11 (12.1) | 12 (18.2) | 0.96 (0.35–2.62) | 0.65 |
| 75+ | 35 (44.3) | 33 (47.1) | 0.73 (0.40–1.34) | |
| **OBESITY** | | | | |
| No | 38 (27.1) | 33 (33.3) | 0.70 (0.38–1.26) | 0.86 |
| Yes | 8 (26.7) | 12 (32.4) | 0.62 (0.18–2.19) | |
| **DIABETES** | | | | |
| No | 34 (29.8) | 31 (33.0) | 0.73 (0.40–1.35) | 0.62 |
| Yes | 12 (21.4) | 14 (33.3) | 0.53 (0.18–1.60) | |
| **HEART FAILURE** | | | | |
| No | 40 (25.5) | 33 (28.5) | 0.81 (0.45–1.44) | 0.15 |
| Yes | 6 (46.2) | 12 (60.0) | 0.24 (0.05–1.09) | |
| **BACKGROUND CV RISK** | | | | |
| CV risks factors | 26 (23.9) | 23 (29.1) | 0.76 (0.37–1.52) | 0.78 |
| CV diseases | 20 (32.8) | 22 (38.6) | 0.65 (0.28–1.51) | |
| **SEVERITY SCORE** | | | | |
| 0 – 3 | 25 (22.9) | 25 (27.5) | 0.62 (0.31–1.25) | 0.59 |
| 4 – 7 | 21 (34.4) | 20 (44.4) | 0.77 (0.36–1.66) | |
| **IN–HOSPITAL USE OF:** | | | | |
| **CORTICOSTEROIDS** | | | | |
| No | 27 (30.7) | 22 (24.2) | 0.88 (0.43–1.82) | 0.16 |
| Yes | 19 (23.2) | 23 (51.1) | 0.40 (0.17–0.90) | |
| **BETA BLOCKERS** | | | | |
| No | 38 (26.4) | 35 (31.0) | 0.74 (0.42–1.32) | 0.58 |
| Yes | 8 (30.8) | 10 (43.5) | 0.50 (0.14–1.79) | |

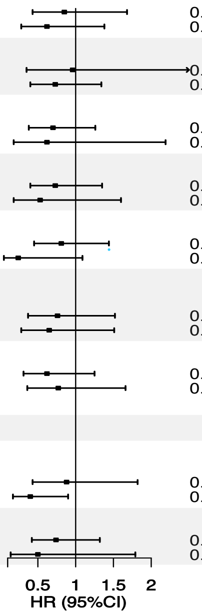


| | ARBs discontinued Deaths (%) | ARBs continued Deaths (%) | MC–HR (95%CI)* | Test of interaction p–value |
|---|---|---|---|---|
| **RISK FACTORS:** | | | | |
| **GENDER** | | | | |
| Females | 17 (25.4) | 8 (14.0) | 2.68 (0.82–8.78) | 0.42 |
| Males | 31 (30.7) | 18 (26.5) | 1.51 (0.74–3.09) | |
| **AGE (years)** | | | | |
| < 75 | 10 (12.7) | 5 (7.9) | 1.43 (0.38–5.30) | 0.84 |
| 75+ | 38 (42.7) | 21 (33.9) | 1.67 (0.84–3.31) | |
| **OBESITY** | | | | |
| No | 36 (25.5) | 19 (22.9) | 1.30 (0.66–2.55) | 0.08 |
| Yes | 12 (44.4) | 7 (16.7) | 5.40 (1.25–23.3) | |
| **DIABETES** | | | | |
| No | 21 (20.8) | 13 (18.8) | 1.35 (0.51–3.57) | 0.74 |
| Yes | 27 (40.3) | 13 (23.2) | 1.67 (0.78–3.61) | |
| **HEART FAILURE** | | | | |
| No | 40 (26.3) | 19 (17.8) | 1.73 (0.88–3.42) | 0.57 |
| Yes | 8 (50.0) | 7 (38.9) | 1.02 (0.19–5.50) | |
| **BACKGROUND CV RISK** | | | | |
| CV risks factors | 23 (20.0) | 12 (15.6) | 1.53 (0.65–3.61) | 0.99 |
| CV diseases | 25 (47.2) | 14 (29.2) | 1.54 (0.64–3.71) | |
| **SEVERITY SCORE** | | | | |
| 0 – 3 | 25 (23.8) | 15 (17.2) | 1.75 (0.79–3.89) | 0.84 |
| 4 – 7 | 23 (36.5) | 11 (29.0) | 1.54 (0.59–4.01) | |
| **IN–HOSPITAL USE OF:** | | | | |
| **CORTICOSTEROIDS** | | | | |
| No | 16 (19.3) | 8 (12.5) | 1.54 (0.48–4.99) | 0.99 |
| Yes | 32 (37.7) | 18 (29.5) | 1.53 (0.77–3.04) | |
| **BETA BLOCKERS** | | | | |
| No | 34 (25.2) | 17 (19.3) | 1.41 (0.69–2.89) | 0.81 |
| Yes | 14 (42.4) | 9 (24.3) | 1.18 (0.34–4.10) | |

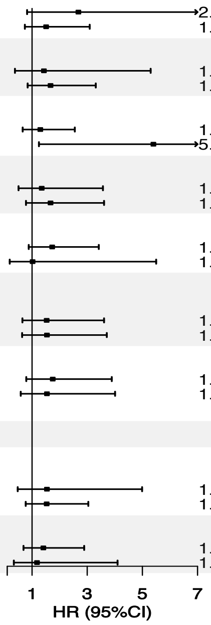


**Abbreviations:** ACEIs: angiotensin-converting enzyme inhibitors; ARBs: angiotensin receptor blockers; MC-HR: mediators-controlled hazard ratio; CV: cardiovascular

* Mediators-controlled hazard ratio (controlled direct effect): including systemic corticosteroids (except when stratification by corticosteroids), anticoagulants and immunomodulators

**Table S1. Characteristics of patients who discontinued vs those who continued with ACEIs.** All were users of ACEIs prior to admission. Baseline comorbidities, outpatient medications and intrahospital treatment

| | ACEIs discontinued n=170 (%) | ACEIs continued n=136 (%) | Standardized difference | p-value |
|---|---|---|---|---|
| **Baseline characteristics** | | | | |
| **Sex,** males | 111 (65.3) | 81 (59.6) | +0.12 | 0.30 |
| **Age,** years, median (IQR) | 73 (63-82) | 75 (68-82) | -0.17 | 0.26 |
| **Hypertension** | 166 (97.6) | 133 (97.8) | 0.01 | 0.93 |
| **Obesity** | 30 (17.6) | 37 (27.2) | -0.23 | **0.05** |
| **Smoking:** | | | | |
| Non-smoker | 61 (35.9) | 54 (39.7) | | |
| Current smoker | 9 (5.3) | 7 (5.1) | - | 0.51 |
| Past smoker | 51 (30.0) | 46 (33.8) | | |
| Not recorded | 49 (28.8) | 29 (21.3) | | |
| **Diabetes** | 56 (32.9) | 42 (30.9) | +0.04 | 0.70 |
| **Dyslipidemia** | 112 (65.9) | 96 (70.6) | -0.10 | 0.38 |
| **CV risk:** | | | | |
| CV risk factors | 109 (64.1) | 79 (58.1) | +0.12 | 0.28 |
| History of CV disease | 61 (35.9) | 57 (41.9) | | |
| **COPD** | 18 (10.6) | 19 (14.0) | -0.10 | 0.37 |
| **Asthma** | 13 (7.6) | 7 (5.1) | +0.10 | 0.38 |
| **Cancer:** | | | | |
| Antecedents | 19 (11.2) | 15 (11.0) | +0.005 | 0.97 |
| Current | 22 (12.9) | 14 (10.3) | +0.08 | 0.48 |
| **Chronic renal failure** | 18 (10.6) | 13 (9.6) | +0.03 | 0.77 |
| **Outpatient medication**: | | | | |
| AMRs | 5 (2.9) | 6 (4.4) | -0.08 | 0.49 |
| CCBs | 50 (29.4) | 26 (19.1) | +0.24 | **0.04** |
| Diuretics | 74 (43.5) | 60 (44.1) | +0.01 | 0.92 |
| Beta-blockers | 39 (22.9) | 28 (20.6) | +0.06 | 0.62 |
| Alpha-blockers | 6 (3.5) | 7 (5.1) | -0.08 | 0.49 |
| Oral anticoagulants | 24 (14.1) | 29 (21.3) | -0.19 | 0.10 |
| Antiplatelet agents | 47 (27.6) | 43 (31.6) | -0.09 | 0.45 |
| NSAIDs | 16 (9.4) | 8 (5.9) | +0.13 | 0.25 |
| Corticosteroids | 15 (8.8) | 3 (2.2) | +0.29 | **0.01** |
| Paracetamol | 87 (51.2) | 81 (59.6) | -0.17 | 0.14 |
| Metamizole | 37 (21.8) | 38 (27.9) | -0.14 | 0.21 |
| Statins | 91 (53.5) | 81 (59.6) | -0.12 | 0.29 |
| Ezetimibe | 8 (4.7) | 3 (2.2) | +0.14 | 0.24 |
| Glucose lowering drugs | 47 (27.6) | 35 (25.7) | +0.04 | 0.71 |
| Insulin | 12 (7.1) | 13 (9.6) | -0.09 | 0.43 |
| **Pneumonia** | 159 (93.5) | 117 (86.0) | +0.25 | **0.03** |

| | | | | |
|---|---|---|---|---|
| **Severity score** | | | | |
| 0-1 | 20 (11.8) | 20 (14.7) | | |
| **2** | 34 (20.0) | 30 (22.1) | | |
| **3** | 55 (32.4) | 41 (30.1) | - | 0.88 |
| **4** | 44 (25.9) | 36 (26.5) | | |
| **5** | 13 (7.6) | 7 (5.1) | | |
| **6-7** | 4 (2.4) | 2 (1.5) | | |
| **Mean (SD)** | 3.0 (1.2) | 2.9 (1.2) | +0.12 | 0.30 |
| **In-hospital stay and treatments received in hospital** | | | | |
| **Stay, days, median (IQR)** | 11 (7-17) | 10 (6-15.5) | +0.05 | 0.23 |
| **ICU admission** | 9 (5.3) | 7 (5.1) | 0.01 | 0.95 |
| **Anticoagulants**: | | | | |
| Oral | 12 (7.1) | 20 (14.7) | -0.25 | **0.03** |
| Parenteral | 136 (80.0) | 79 (58.1) | +0.49 | **<0.001** |
| **Antiplatelet drugs** | 43 (25.3) | 43 (31.6) | -0.14 | 0.22 |
| **Statins** | 27 (15.9) | 59 (43.4) | -0.63 | **<0.001** |
| **Glucose lowering drugs**: | | | | |
| Oral | 7 (4.1) | 19 (14.0) | -0.35 | **0.002** |
| Insulin | 56 (32.9) | 38 (27.9) | +0.11 | 0.35 |
| **Hydroxychloroquine** | 151 (88.8) | 115 (84.6) | +0.13 | 0.27 |
| **Lopinavir+Ritonavir/ Darunavir+Cobicistat** | 142 (83.5) | 109 (80.1) | +0.09 | 0.44 |
| **Azithromycin** | 78 (45.9) | 52 (38.2) | +0.15 | 0.18 |
| **Other macrolides** | 3 (1.8) | 7 (5.1) | -0.19 | 0.10 |
| **Other antivirals*** | 3 (1.8) | 5 (3.7) | -0.12 | 0.30 |
| **Other antibacterial agents** | 113 (66.5) | 92 (67.6) | -0.02 | 0.83 |
| **Immunomodulating agents**: | | | | |
| Tocilizumab | 22 (12.9) | 30 (22.1) | -0.24 | **0.04** |
| Others** | 53 (31.2) | 59 (43.4) | -0.25 | **0.03** |
| **Corticosteroids** | 82 (48.2) | 45 (33.1) | +0.31 | **0.01** |
| **Antihypertensive drugs**: | | | | |
| CCBs | 66 (38.8) | 29 (21.3) | +0.39 | **0.001** |
| Beta-blockers | 26 (15.3) | 23 (16.9) | -0.04 | 0.70 |
| Low-ceiling diuretics | 6 (3.5) | 14 (10.3) | -0.27 | **0.02** |
| High-ceiling diuretics | 26 (15.3) | 22 (16.2) | +0.02 | 0.83 |
| Alpha-blockers | 2 (1.2) | 3 (2.2) | -0.08 | 0.48 |

**Abbreviations:** ACEIs: angiotensin-converting enzyme inhibitors; IQR: interquartile range; CV: cardiovascular; COPD: chronic obstructive pulmonary disease; AMRs: antagonists of mineralocorticoid receptor; CCBs: calcium channel blockers NSAIDs: nonsteroidal anti-inflammatory drugs; SD: standard deviation; ICU: intensive care unit

* Other antivirals: remdesivir, aciclovir, bictegravir-emtricitabine-tenofovir, tenofovir, emtricitabine-tenofovir, lamivudine-abacabir-dolutegravir, valaciclovir and valganciclovir.

** Other immunomodulators: Jak inhibitors, interferon beta-1b, ciclosporin, anakinra, ceftriaxone, leflunomide, methotrexate and mycophenolic acid.

**Table S2. Characteristics of patients who discontinued vs those who continued with ARBs.** All were users of ARBs prior to admission. Baseline comorbidities, outpatient medications and intrahospital treatment

| | ARBs discontinued n=168 (%) | ARBs continued n=125 (%) | Standardized difference | p-value |
|---|---|---|---|---|
| **Baseline characteristics** | | | | |
| **Sex,** males | 101 (60.1) | 68 (54.4) | +0.12 | 0.33 |
| **Age,** years, median (IQR) | 75 (67-82) | 74 (69-82) | +0.12 | 0.74 |
| **Hypertension** | 164 (97.6) | 120 (96.0) | +0.09 | 0.43 |
| **Obesity** | 27 (16.1) | 42 (33.6) | -0.41 | **<0.001** |
| **Smoking:** | | | | |
| Non-smoker | 66 (39.3) | 52 (41.6) | | |
| Current smoker | 5 (3.0) | 6 (4.8) | | |
| Past smoker | 51 (30.4) | 30 (24.0) | - | 0.59 |
| Not recorded | 46 (27.4) | 37 (29.6) | | |
| **Diabetes** | 67 (39.9) | 56 (44.8) | -0.10 | 0.40 |
| **Dyslipidemia** | 106 (63.1) | 80 (64.0) | -0.02 | 0.87 |
| **CV risk:** | | | | |
| CV risk factors | 115 (68.5) | 77 (61.6) | +0.14 | 0.22 |
| **History of CV disease** | 53 (31.5) | 48 (38.4) | | |
| **COPD** | 24 (14.3) | 13 (10.4) | +0.12 | 0.32 |
| **Asthma** | 14 (8.3) | 11 (8.8) | +0.02 | 0.89 |
| **Cancer:** | | | | |
| Antecedents | 13 (7.7) | 13 (10.4) | -0.09 | 0.43 |
| Current | 13 (7.7) | 12 (9.6) | +0.07 | 0.57 |
| **Chronic Renal Failure** | 20 (11.9) | 18 (14.4) | -0.07 | 0.53 |
| **Baseline co-medication**: | | | | |
| AMRs | 6 (3.6) | 6 (4.8) | -0.06 | 0.60 |
| CCBs | 64 (38.1) | 48 (38.4) | -0.01 | 0.96 |
| Diuretics | 98 (58.3) | 80 (64.0) | -0.12 | 0.33 |
| Beta-blockers | 41 (24.4) | 37 (29.6) | -0.12 | 0.32 |
| Alpha-blockers | 14 (8.3) | 12 (9.6) | -0.04 | 0.71 |
| Oral anticoagulants | 31 (18.5) | 22 (17.6) | +0.02 | 0.85 |
| Antiplatelet agents | 45 (26.8) | 33 (26.4) | +0.01 | 0.94 |
| NSAIDs | 13 (7.7) | 9 (7.2) | +0.02 | 0.86 |
| Corticosteroids | 10 (6.0) | 7 (5.6) | +0.02 | 0.90 |
| Paracetamol | 89 (53.0) | 80 (64.0) | -0.22 | 0.06 |
| Metamizole | 57 (33.9) | 33 (26.4) | +0.16 | 0.17 |
| Statins | 81 (48.2) | 69 (55.2) | -0.14 | 0.24 |
| Ezetimibe | 6 (3.6) | 9 (7.2) | +0.16 | 0.16 |
| Glucose lowering drugs | 53 (31.5) | 50 (40.0) | -0.18 | 0.13 |
| Insulin | 21 (11.9) | 20 (16.0) | -0.12 | 0.31 |
| **Pneumonia** | 158 (94.1) | 115 (92.0) | +0.09 | 0.49 |

| | | | | |
|---|---|---|---|---|
| **Severity score** | | | | |
| 0-1 | 19 (11.3) | 15 (12.0) | | |
| 2 | 32 (19.0) | 42 (33.6) | - | 0.05 |
| 3 | 54 (32.1) | 30 (24.0) | | |
| 4 | 34 (20.2) | 26 (20.8) | | |
| 5 | 26 (15.5) | 10 (8.0) | | |
| 6-7 | 3 (1.8) | 2 (1.6) | | |
| Mean (SD) | 3.2 (1.3) | 2.8 (1.2) | +0.25 | **0.03** |
| **In-hospital stay and treatments received in hospital** | | | | |
| **Stay,** days, median (IQR) | 11 (8-17) | 12 (7-16) | -0.04 | 0.79 |
| **ICU admission** | 10 (6.0) | 9 (7.2) | +0.05 | 0.67 |
| **Anticoagulants**: | | | | |
| Oral | 14 (8.3) | 16 (12.8) | -0.15 | 0.21 |
| Parenteral | 135 (80.4) | 69 (55.2) | +0.56 | **<0.001** |
| **Antiplatelet drugs** | 36 (21.4) | 37 (29.6) | -0.19 | 0.11 |
| **Statins** | 19 (11.3) | 50 (40.0) | -0.69 | **<0.001** |
| **Glucose lowering drugs**: | | | | |
| Oral | 7 (4.2) | 22 (17.6) | -0.44 | **<0.001** |
| Insulin | 69 (41.1) | 40 (32.0) | +0.19 | 0.11 |
| **Hydroxychloroquine** | 153 (91.1) | 108 (86.4) | +0.15 | 0.20 |
| **Lopinavir+Ritonavir/ Darunavir+Cobicistat** | 143 (85.1) | 104 (83.2) | +0.05 | 0.66 |
| **Azithromycin** | 50 (29.8) | 52 (41.6) | -0.25 | **0.04** |
| **Other macrolides** | 7 (4.2) | 10 (8.0) | -0.16 | 0.17 |
| **Other antivirals*** | 5 (3.0) | 1 (0.8) | +0.16 | 0.19 |
| **Other antibacterial agents** | 98 (58.3) | 85 (68.0) | -0.20 | 0.09 |
| **Immunomodulating agents**: | | | | |
| Tocilizumab | 21 (12.5) | 23 (18.4) | -0.16 | 0.16 |
| Others** | 45 (26.8) | 54 (43.2) | -0.35 | **0.003** |
| **Corticosteroids** | 85 (50.6) | 61 (48.8) | +0.04 | 0.76 |
| **Antihypertensive drugs**: | | | | |
| CCBs | 64 (38.1) | 36 (28.8) | +0.20 | 0.10 |
| Beta-blockers | 33 (19.6) | 37 (29.6) | -0.23 | 0.05 |
| Low-ceiling diuretics | 11 (6.5) | 33 (26.4) | -0.55 | **<0.001** |
| High-ceiling diuretics | 25 (14.9) | 14 (11.2) | +0.11 | 0.36 |
| Alpha-blockers | 9 (5.4) | 6 (4.8) | +0.03 | 0.83 |

**Abbreviations:** ACEIs: angiotensin-converting enzyme inhibitors; IQR: interquartile range; CV: cardiovascular; COPD: chronic obstructive pulmonary disease; AMRs: antagonists of mineralocorticoid receptor; CCBs: calcium channel blockers NSAIDs: nonsteroidal anti-inflammatory drugs; SD: standard deviation; ICU: intensive care unit

* Other antivirals: remdesivir, aciclovir, bictegravir-emtricitabine-tenofovir, tenofovir, emtricitabine-tenofovir, lamivudine-abacabir-dolutegravir, valaciclovir and valganciclovir.

** Other immunomodulators: Jak inhibitors, interferon beta-1b, ciclosporin, anakinra, ceftriaxone, leflunomide, methotrexate and mycophenolic acid.

**Table S3. Characteristics of patients who continued with ACEIs as compared to those who continued with ARBs.** All patients were prior users of ACEIs or ARBs. Baseline comorbidities, outpatient medication and intrahospital treatment.

| | ACEIs continued n=136 (%) | ARBs continued n=125 (%) | Standardized difference | p-value |
|---|---|---|---|---|
| **Baseline characteristics** | | | | |
| **Sex, m**ales | 81 (59.6) | 68 (54.4) | +0.10 | 0.40 |
| **Age,** years, median (IQR) | 75 (68-82) | 74 (69-82) | +0.09 | 0.89 |
| **Hypertension** | 133 (97.8) | 120 (96.0) | +0.10 | 0.40 |
| **Obesity** | 37 (27.2) | 42 (33.6) | -0.14 | 0.26 |
| **Smoking:** | | | | |
| Non-smoker | 54 (39.7) | 52 (41.6) | | |
| Current smoker | 7 (5.1) | 6 (4.8) | - | 0.26 |
| Past smoker | 46 (33.8) | 30 (24.0) | | |
| Not recorded | 29 (21.3) | 37 (29.6) | | |
| **Diabetes** | 42 (30.9) | 56 (44.8) | -0.29 | **0.02** |
| **Dyslipidemia** | 96 (70.6) | 80 (64.0) | +0.14 | 0.26 |
| **CV risk:** | | | | |
| CV risk factors | 79 (58.1) | 77 (61.6) | -0.07 | 0.56 |
| History of CV disease | 57 (41.9) | 48 (38.4) | | |
| **COPD** | 19 (14.0) | 13 (10.4) | +0.11 | 0.38 |
| **Asthma** | 7 (5.1) | 11 (8.8) | -0.14 | 0.24 |
| **Cancer:** | | | | |
| Antecedents | 15 (11.0) | 13 (10.4) | +0.02 | 0.87 |
| Current | 14 (10.3) | 12 (9.6) | +0.02 | 0.85 |
| **Chronic Renal Failure** | 13 (9.6) | 18 (14.4) | -0.15 | 0.23 |
| **Medication before admission**: | | | | |
| AMRs | 6 (4.4) | 6 (4.8) | -0.02 | 0.88 |
| CCBs | 26 (19.1) | 48 (38.4) | -0.43 | **<0.001** |
| Diuretics | 60 (44.1) | 80 (64.0) | -0.41 | **0.001** |
| Beta-blockers | 28 (20.6) | 37 (29.6) | -0.21 | 0.09 |
| Alpha-blockers | 7 (5.1) | 12 (9.6) | -0.17 | 0.17 |
| Oral anticoagulants | 29 (21.3) | 22 (17.6) | +0.09 | 0.45 |
| Antiplatelet agents | 43 (31.6) | 33 (26.4) | +0.11 | 0.35 |
| NSAIDs | 8 (5.8) | 9 (7.1) | -0.05 | 0.65 |
| Corticosteroids | 3 (2.2) | 7 (5.6) | -0.18 | 0.15 |
| Paracetamol | 81 (59.6) | 80 (64.0) | -0.09 | 0.46 |
| Metamizole | 38 (27.9) | 33 (26.4) | +0.03 | 0.78 |
| Statins | 81 (59.6) | 69 (55.2) | +0.09 | 0.48 |
| Ezetimibe | 3 (2.2) | 9 (7.2) | -0.24 | 0.05 |
| Glucose lowering drugs | 35 (25.7) | 50 (40.0) | -0.31 | **0.01** |
| Insulin | 13 (9.6) | 20 (16.0) | -0.19 | 0.12 |
| **Pneumonia** | 117 (86.0) | 115 (92.0) | -0.19 | 0.13 |

| | | | | |
|---|---|---|---|---|
| **Severity score** | 20 (14.7) | 15 (12.0) | | |
| 0-1 | 30 (22.1) | 42 (33.6) | | |
| 2 | 41 (30.1) | 30 (24.0) | | |
| 3 | 36 (26.5) | 26 (20.8) | - | 0.30 |
| 4 | 7 (5.1) | 10 (8.0) | | |
| 5 | 2 (1.5) | 2 (1.6) | | |
| 6-7 | 2.9 (1.2) | 2.8 (1.2) | 0.05 | 0.66 |
| Mean (SD) | | | | |
| **In-hospital stay and treatments received in hospital** | | | | |
| **Stay,** days, median (IQR) | 10 (6-15.5) | 12 (7-16) | -0.14 | 0.17 |
| **ICU admission** | 7 (5.1) | 9 (7.2) | -0.09 | 0.49 |
| **Anticoagulants**: | | | | |
| Oral | 20 (14.7) | 16 (12.8) | +0.06 | 0.66 |
| Parenteral | 79 (58.1) | 69 (55.2) | +0.06 | 0.64 |
| **Antiplatelet drugs** | 43 (31.6) | 37 (29.6) | +0.04 | 0.72 |
| **Statins** | 59 (43.4) | 50 (40.0) | +0.07 | 0.58 |
| **Glucose lowering drugs:** | | | | |
| Oral | 19 (14.0) | 22 (17.6) | -0.10 | 0.42 |
| Insulin | 38 (27.9) | 40 (32.0) | +0.09 | 0.47 |
| **Hydroxychloroquine** | 115 (84.6) | 108 (86.4) | -0.05 | 0.67 |
| **Lopinavir+Ritonavir/ Darunavir+Cobicistat** | 109 (80.1) | 104 (83.2) | -0.08 | 0.52 |
| **Azithromycin** | 52 (38.2) | 52 (41.6) | -0.07 | 0.58 |
| **Other macrolides** | 7 (5.1) | 10 (8.0) | -0.11 | 0.35 |
| **Other antivirals*** | 5 (3.7) | 1 (0.8) | +0.19 | 0.12 |
| **Other antibacterial agents** | 92 (67.6) | 85 (68.0) | 0.01 | 0.95 |
| **Immunomodulating agents**: | | | | |
| Tocilizumab | 30 (22.1) | 23 (18.4) | +0.09 | 0.46 |
| Others** | 59 (43.4) | 54 (43.2) | +0.004 | 0.98 |
| **Corticosteroids** | 45 (33.1) | 61 (48.8) | -0.32 | **0.01** |
| **Antihypertensive drugs**: | | | | |
| CCBs | 29 (21.3) | 36 (28.8) | -0.17 | 0.16 |
| Beta-blockers | 23 (16.9) | 37 (29.6) | -0.30 | **0.02** |
| Low-ceiling diuretics | 14 (10.3) | 33 (26.4) | -0.42 | **<0.001** |
| High-ceiling diuretics | 22 (16.2) | 14 (11.2) | +0.14 | 0.24 |
| Alpha-blockers | 3 (2.2) | 6 (4.8) | -0.14 | 0.25 |

**Abbreviations:** ACEIs: angiotensin-converting enzyme inhibitors; ARBs: angiotensin receptor blockers; IQR: interquartile range; CV: cardiovascular; COPD: chronic obstructive pulmonary disease; AMRs: antagonists of mineralocorticoid receptor; CCBs: calcium channel blockers NSAIDs: nonsteroidal anti-inflammatory drugs; SD: standard deviation; ICU: intensive care unit

* Other antivirals: remdesivir, aciclovir, bictegravir-emtricitabine-tenofovir, tenofovir, emtricitabine-tenofovir, lamivudine-abacabir-dolutegravir, valaciclovir and valganciclovir.

** Other immunomodulators: Jak inhibitors, interferon beta-1b, ciclosporin, anakinra, ceftriaxone, leflunomide, methotrexate and mycophenolic acid.

**Table S4: Sensitivity analyses**

***1. Reclassification of uncertain to RASIs discontinued if they had a sole prescription on day 1 and to RASIs continued if they had a sole prescription on days 2 or 3.***

| Outcome | RASIs discontinued N=362 | | RASIs continued N=316 | | Crude HR (95%CI) | PS-adj HR* (95%CI) | MC-HR** (95%CI) |
|---|---|---|---|---|---|---|---|
| | Patients with event | Event rate (%) | Patients with event | Event rate (%) | | | |
| **Death** | 100 | 27.6 | 84 | 26.6 | 1.02 (0.77-1.37) | 1.15 (0.82-1.60) | 1.16 (0.83-1.62) |
| **Death + ICU** | 115 | 31.8 | 95 | 30.1 | 1.03 (0.79-1.35) | 1.17 (0.86-1.61) | 1.22 (0.89-1.67) |

***2. Reclassification of all uncertain to the cohort of patients who discontinued***

| Outcome | RASIs discontinued N=393 | | RASIs continued N=285 | | Crude HR (95%CI) | PS-adj HR* (95%CI) | MC-HR** (95%CI) |
|---|---|---|---|---|---|---|---|
| | Patients with event | Event rate (%) | Patients with event | Event rate (%) | | | |
| **Death** | 105 | 26.7 | 79 | 27.7 | 0.90 (0.68-1.21) | 0.98 (0.69-1.39) | 0.99 (0.70-1.41) |
| **Death + ICU** | 121 | 30.8 | 89 | 31.2 | 0.93 (0.70-1.22) | 1.02 (0.74-1.42) | 1.06 (0.76-1.48) |

***3. Using a 2-days window (uncertain excluded)***

| Outcome | RASIs discontinued N=358 | | RASIs continued N=227 | | Crude HR (95%CI) | PS-adj HR* (95%CI) | MC-HR** (95%CI) |
|---|---|---|---|---|---|---|---|
| | Patients with event | Event rate (%) | Patients with event | Event rate (%) | | | |
| **Death** | 98 | 27.4 | 65 | 28.6 | 0.90 (0.66-1.23) | 0.87 (0.59-1.30) | 0.90 (0.60-1.35) |
| **Death + ICU** | 112 | 31.3 | 73 | 32.2 | 0.91 (0.68-1.22) | 0.93 (0.63-1.35) | 0.96 (0.66-1.40) |

***4. Using a 2-days window (uncertain reclassified to RASIs discontinued if they had a sole prescription on day 1 and to RASIs continued if had a sole prescription on day 2)***

| Outcome | RASIs discontinued N=380 | | RASIs continued N=298 | | Crude HR (95%CI) | PS-adj HR* (95%CI) | MC-HR** (95%CI) |
|---|---|---|---|---|---|---|---|
| | Patients with event | Event rate (%) | Patients with event | Event rate (%) | | | |
| **Death** | 104 | 27.4 | 80 | 26.9 | 0.97 (0.73-1.30) | 1.09 (0.77-1.54) | 1.10 (0.78-1.56) |
| **Death + ICU** | 120 | 31.6 | 90 | 30.2 | 1.01 (0.77-1.32) | 1.15 (0.83-1.59) | 1.17 (0.85-1.62) |

**Abbreviations:** RASIs: renin-angiotensin system inhibitors; HR: hazard ratio; ICU: intensive care unit
*Propensity-scores-adjusted hazard ratio (adjusted total effect)
** Mediators-controlled hazard ratio (controlled direct effect): a) systemic corticosteroids, anticoagulants and immunomodulators when the outcome was death; and b) immunomodulators and anticoagulants when the outcome was death plus ICU admission

**Table S5: Published studies reporting mortality rates associated with inpatient use of renin-angiotensin system inhibitors (RASI)**

| First Author | Country | Definition of inpatient RASI use (or cont) | Primary outcome | Number of inpatients | Disc rate | Main results Adjusted HR/OR (95%CI) |
|---|---|---|---|---|---|---|
| Cannata [9] | Italy | At the time of hospital admission | Mortality | 56 cont RASIs<br>117 disc RASIs<br>224 no RASIs | 67.7% | RASI cont vs disc: 0.17 (0.03-1.00)<br>RASI cont vs no RASI + disc: **0.14 (0.03-0.66)** |
| Chaudhri [12] | US | Any time during hospitalization | In-hospital mortality | 49 cont RASIs<br>31 disc RASIs | 38.8% | RASI cont vs disc: 0.31 (0.08-1.26) |
| Chen [32] | China | Any time during hospitalization | In-hospital mortality | 93 RASIs<br>2530 no RASIs<br>53 RASIs<br>315 non-RASIs | Not reported | RASI vs. no RASI: 0.40 (0.15-1.03)<br>RASI vs. non-RASI: **0.18 (0.04-0.86)** |
| Di Castelnuovo (CORIST) [28] | Italy | Any time during hospitalization | In-hospital mortality | 549 ACEIs<br>542 ARBs<br>2807 no RASIs | 12.4% | ACEIs vs no RASI: 0.96 (0.77-1.20);<br>ARBs vs. no RASI: 0.89 (0.71-1.12) |
| Lahens [17] | France | Previous users who continued during the first 7 days of hospital admission | 30-day mortality | 84 RASIs<br>263 no RASIs | 33.0% | RASI vs non-RASI: **0.25 (0.09-0.65)** |
| Lam [10] | US | Any time during hospitalization | In-hospital mortality | 164 cont RASIs<br>171 disc RASIs | 51.0% | RASI cont vs disc: **0.215 (0.101-0.455)** |
| Li [27] | China | Use at time of admission that continued through hospitalization | In-hospital mortality | 115 RASIs<br>247 non-RASIs | Not reported | HR/OR not reported<br>RASI: 18.3%<br>No-RASI: 22.7% |
| Meng [34] | China | Any time during hospitalization | In-hospital mortality | 17 RASIs<br>25 non-RASIs | Not reported | HR/OR not reported<br>RASI: 0.0%<br>No-RASI: 4.0% |
| Rodilla [30] | Spain | Any time during hospitalization | In-hospital mortality | ACEIs: 490<br>ARBs: 1046 | ACEIs: 55.1%<br>ARBs: 53.6% | In-hospital ACEIs vs non-use: **0.6 (0.45-0.66)**<br>In-hospital ARBs vs non-use: **0.5 (0.45-0.65)** |
| Soleimani [31] | Iran | During 7 days after admission | In-hospital mortality | ARBs:<br>79 cont<br>43 disc | 35.2% | HR/OR not reported<br>ARB cont: 12.7%<br>ARB disc: 53.5% |
| Wang [29] | China | Any time during hospitalization | Mortality | 81 RASIs<br>129 non-RASIs | Not reported | HR/OR not reported<br>RASI: 8.64%<br>Non-RASI: 3.88% |
| Zhang [11] | China | Any time during hospitalization | 28-day all-cause mortality | 188 RASIs<br>940 no RASIs | Not reported | RASI vs non-RASI: **0.42 (0.19-0.92)** |
| Zhou [33] | China | Any time during hospitalization | 28-day all-cause mortality | 989 RASIs<br>2583 no RASIs | Not reported | RASI vs non-RASI: **0.39 (0.26-0.58)** |

**Abbreviations:** ACEIs: angiotensin converting enzyme inhibitors; ARBs: angiotensin receptor blocker; Cont: continuation; Disc: discontinuation; Disc rate: discontinuation rate; HR: hazard ratio; ICU: intensive care unit; OR: odds ratio; RASIs: renin-angiotensin system inhibitors
**Note:** In this table, "non-RASIs" means that patients received other antihypertensive drugs different from RASIs, while "no RASIs" means that patients were not exposed to RASIs (but not necessarily exposed to other antihypertensive drugs). Statistically significant HR/OR values are in bold.
